# Modelling how nutritional solutions may help alleviate paediatric iron deficiency: Application to the Indian subcontinent

**DOI:** 10.64898/2026.01.02.26343339

**Authors:** Bharadwaj Vemparala, Shreya Chowdhury, Akshay Tiwari, Venu Prasad S, Kaushiki Prabhudesai, Karan Lomore, Kruti Shah, Preethi Rahul, Priya Karkera, Narendra M. Dixit, K. V. Venkatesh

## Abstract

Iron deficiency (ID) in children caused by dietary deprivation can be alleviated in ∼40% cases with iron-fortified nutritional solutions. Devising more effective nutritional solutions requires a quantitative understanding of iron homeostasis, which is lacking. Here, we developed a physiologically-based mathematical model of paediatric iron homeostasis that recapitulates ID due to dietary deprivation and its alleviation using nutritional solutions. The model integrates key cellular and systemic iron regulatory processes, associated anatomical sites, the gut microbiota, and nutritional solutions, and shows how their complex interplay governs iron homeostasis. The model quantitatively described the alleviation of ID with nutritional solutions in a large clinical trial in Bangladesh. In virtual Indian populations, the model predicts that over-the-counter iron-fortified nutritional solutions can restore haemoglobin level to normalcy in >85% mildly anaemic infants and young children. Our study thus elucidates individual- and population-level underpinnings of ID, describes clinical observations, and informs efforts to improve intervention strategies.

## Introduction

Iron deficiency (ID) is the most prevalent nutrient deficiency in children^1^. The resulting anaemia, affecting ∼40% children globally^1^, has long-term developmental consequences, creating significant disease burden, particularly in low- and middle-income countries like India^2–4^. Among the major causes of ID are inadequate dietary supply of iron and its poor absorption, worsened by compromised gut health and/or inflammation^5^. WHO estimates that about 42% of the ID cases can be alleviated by iron supplementation, offering an important public health intervention^5^. It would be of great benefit to improve the outcomes of such interventions, addressing the large gap beyond the modest ∼42% success rate today. A key knowledge gap limiting such improvements is the lack of a quantitative understanding of the mechanisms governing iron homeostasis.

Iron is needed for several critical processes in the body. It is a key component of the oxygen-carrying proteins haemoglobin and myoglobin^6^ and is essential to many cellular functions, such as enzymatic processes, DNA synthesis and repair, and mitochondrial energy generation^7^. ID in children thus compromises their health significantly, including their cognitive development^8^. At the same time, non-absorbed iron can favour the growth of pathogenic gut bacteria and trigger the production of toxic reactive oxygen species^9^.

Excess iron may have long-term adverse consequences^10,11^. Thus, iron levels are tightly regulated in the body^5,12^. Broadly, the regulation happens at two distinct but interconnected levels: cellular and systemic^12^. At the cellular level, dietary iron is absorbed from the duodenum by enterocytes, which act also as stores of iron and its exporters to circulation^7^. This iron uptake from the gut lumen is regulated by the amount of iron stored in the enterocytes, with greater stores limiting uptake. This mechanism controls iron uptake at the source and is important to prevent iron overload because our body lacks specialized excretion mechanisms for iron^12^. Systemic regulation is based on the level of iron present in circulation and is governed by a key liver hormone, hepcidin^13,14^. Liver cells also act as stores of iron. Iron is typically stored in cells in ferritin cages. Excess stored iron in the liver cells triggers the secretion of hepcidin, which blocks the export of iron from enterocytes and other cellular stores into circulation^12^. These broad regulatory mechanisms are modulated by several other processes, including RBC synthesis and senescence, competitive growth of commensal and pathogenic gut bacteria, and dietary inputs^12^.

While the different regulatory mechanisms are well understood individually, a systems-level view that recognizes their simultaneous and integrated functioning and offers a quantitative description of iron homeostasis is lacking^14^. A mathematical model has been developed to describe adult iron homeostasis, especially in the context of cancer chemotherapy and chronic kidney disease^11^. Its applicability to dietary deprivation, however, remains unclear given that it does not consider the role of the gut microbiome. Importantly, no models exist for children. Nutritional solutions to address ID in children have therefore remained empirical. Here, to overcome this limitation, we developed a comprehensive, physiologically based, systems-level computational model of paediatric iron homeostasis. We applied it to describe ID and the impact of nutritional solutions at the individual and population levels.

## Results

### Overview of the computational model

Our model provides a quantitative link between the dietary supply of iron and the resulting level of serum haemoglobin so that the effect of dietary deprivation and/or nutritional solution on ID can be predicted. Constructing the model required integrating key regulatory processes underlying iron homeostasis. We present an overview here (Fig. 1, Table S1). Details are in Methods.

**Fig. 1.**
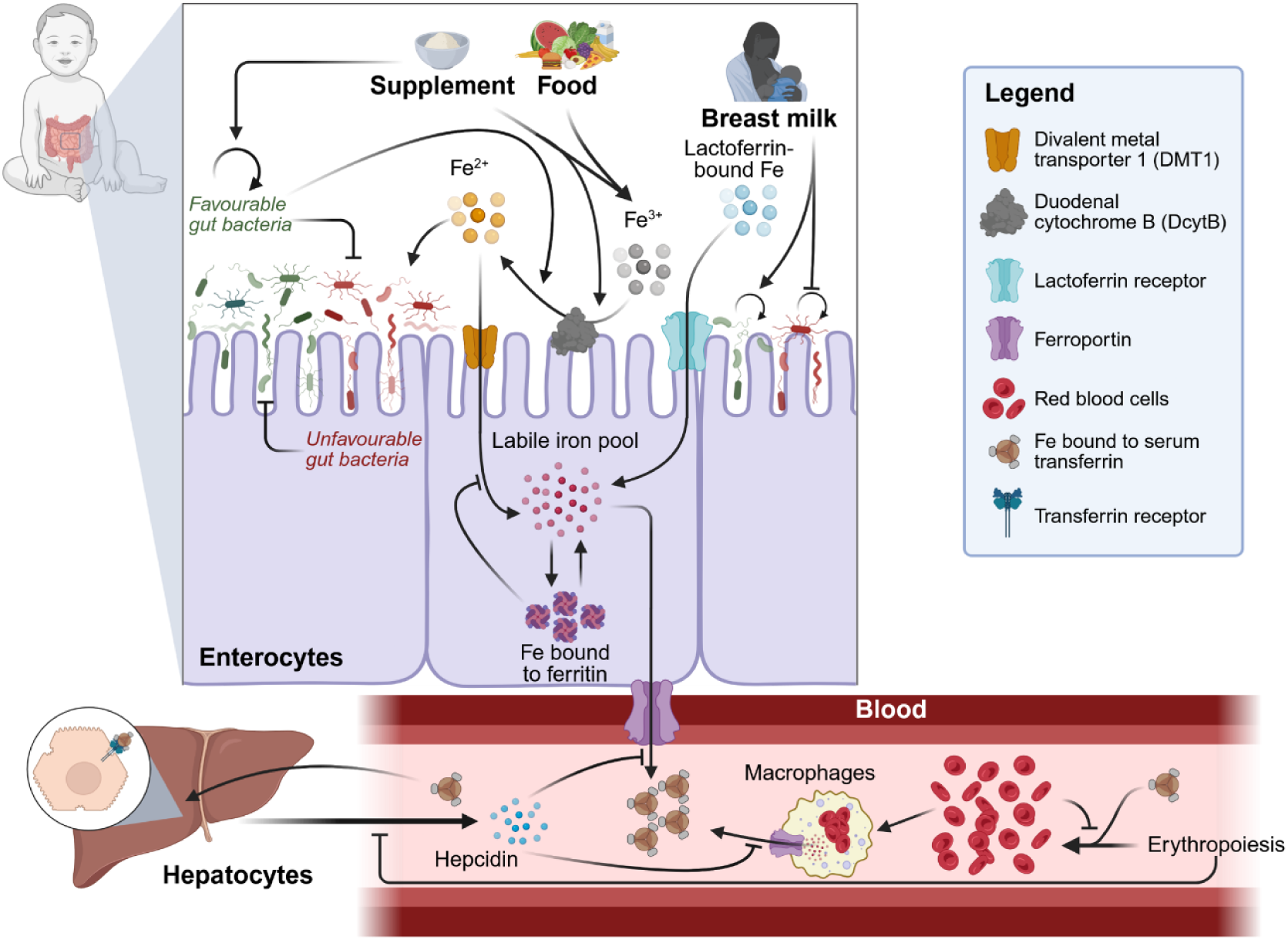
Schematic of the model. Iron from dietary sources is absorbed by enterocytes and transported into circulation. The key processes and molecular players involved in iron uptake, transport, storage, and regulation are displayed. See text for details. Image was created in BioRender (https://www.biorender.com/).

We consider iron sourced from dietary inputs, which includes breast milk and other food sources depending on the age of the children. Breast milk contains iron bound to lactoferrin, which is directly taken up by enterocytes via specialized lactoferrin receptors^15,16^. Other foods, including nutritional solutions, typically contain iron in its oxidized state (Fe^3+^) that is not bound to lactoferrin^17^. The enzyme duodenal cytochrome B (DcytB) on the apical surface of the enterocytes converts iron to its reduced state (Fe^2+^) in a pH-dependent manner^12^. Commensal gut microbes help lower gut pH, favouring iron reduction^18^. Breast milk contains human milk oligosaccharides (HMO), which favour commensal and hinder pathogenic bacterial growth^19,20^. Importantly, iron can be taken up by unfavourable, pathogenic bacteria, which can disturb the balance of the gut microbiota, compromising iron uptake by enterocytes. Iron in its reduced state is absorbed by enterocytes through the divalent metal transporter 1 (DMT1) protein, expressed on enterocytes^12^. The latter uptake is negatively regulated by iron stores in the enterocytes^12^.

Within enterocytes, iron is bound reversibly to and stored in ferritin cages^7^. Free iron forms the labile iron pool (LIP), from which iron is used for cellular metabolic requirements. Iron from the LIP is also bound to ferroportin molecules on the basolateral membrane of the enterocytes and exported into circulation^21^. Hephaestin molecules, colocalized with ferroportin on the membrane^7^, convert iron to its oxidized state at the time of its entry into circulation.

In circulation, iron is bound to transferrin molecules, which are typically in large excess so that free iron is absent in circulation barring in iron overload scenarios^5^. Transferrin-bound iron is primarily trafficked to the bone marrow to be incorporated into haemoglobin molecules during RBC production^5^. RBC levels are under autoregulation: lower RBC levels lead to increased production of the hormone erythropoietin, which in turn increases RBC production^12^. At the same time, the hormone erythroferrone produced during RBC synthesis can downregulate hepcidin, increasing iron export into circulation, and lower erythropoietin production, inhibiting RBC synthesis^12,22^. Senescent RBCs are taken up by macrophages. Iron in these RBCs can be released back into circulation via the ferroportin receptor expressed on the macrophage surfaces^12,23^.

Transferrin-bound iron is also taken up by hepatocytes via transferrin receptor–mediated endocytosis^24^. Internalized iron is stored in ferritin cages and can be secreted out by ferroportin molecules, like with enterocytes. Importantly, the liver also produces the hormone hepcidin, which is a global regulator of circulating iron levels. Hepcidin production is increased when iron levels are high. Hepcidin blocks iron export by ferroportin from hepatocytes, enterocytes, and macrophages, thus regulating circulating iron levels^12,14^. These processes above collectively maintain iron homeostasis in healthy children.

With deprivation of dietary iron, homeostasis can be lost, leading to ID. Nutritional solutions to address this ID are designed to help iron absorption and contain iron in its oxidized state, prebiotics like fructo-oligosaccharides (FOS) and inulin, and/or vitamin C. The iron in these feeds is taken up by enterocytes following the processes above. FOS and inulin preferentially enable the growth of favourable, commensal bacteria and enhance iron uptake^25^. Vitamin C further aids in this uptake in a dose-dependent manner; DcytB is an oxidoreductase that reduces Fe^3+^ and is aided in the process by an electron-acceptor like vitamin C^26,27^.

We constructed equations to quantify the above processes and describe how haemoglobin levels are maintained under healthy conditions, the extent to which they are compromised under dietary deprivation, and the level of their restoration with nutritional solutions designed to foster iron absorption (Methods). We examined population-level effects by generating virtual populations based on known variations in these parameters. We assessed effects in three age groups: 4-months, 18-months, and 24-months (Table S2), the first representing children exclusively on breast milk, the second receiving a mix of breast milk and solid food, and the third exclusively on solid food.

### Iron homeostasis in healthy children and the effect of dietary deprivation

We considered first a 4-month-old infant of median weight, exclusively on breast milk (Table S2), at the recommended level of 800 mL breast milk in ∼12 feeds per day^28^. Milk from a healthy mother contains ∼10 g/L of HMO^29^ and ∼0.3 mg/L of iron bound to lactoferrin, with half of the latter bioavailable^30^. Under these feeding conditions and with representative model parameter values (Table 1), our model predicted serum haemoglobin levels in the normal range (Fig. S1). It also predicted normal levels of other key entities involved in iron homeostasis, namely transferrin-bound iron, hepcidin, RBC, and the extent of favourable and unfavourable bacteria in the gut microbiome (Fig. S1), recapitulating the state of a healthy child.

We next simulated the effect of dietary deprivation, due to lower volume of breast milk per feed (Fig. 2a-b, Fig. S2) or lower frequency of feeding (Fig. 2c-d, Fig. S3). HMO and lactoferrin-bound iron levels were predicted to peak at lower values with lower feed volumes (Fig. 2a). This resulted in lower haemoglobin levels (Fig. 2b). For instance, with 50% feed volume, haemoglobin levels dropped to ∼8.7 g/dL from the normal level of ∼11.3 g/dL, indicating anaemia (Fig. 2b). Similarly, with feeding frequency reduced to 8 per day, haemoglobin levels dropped to ∼9.8 g/dL, again indicating anaemia (Fig. 2d).

**Fig. 2.**
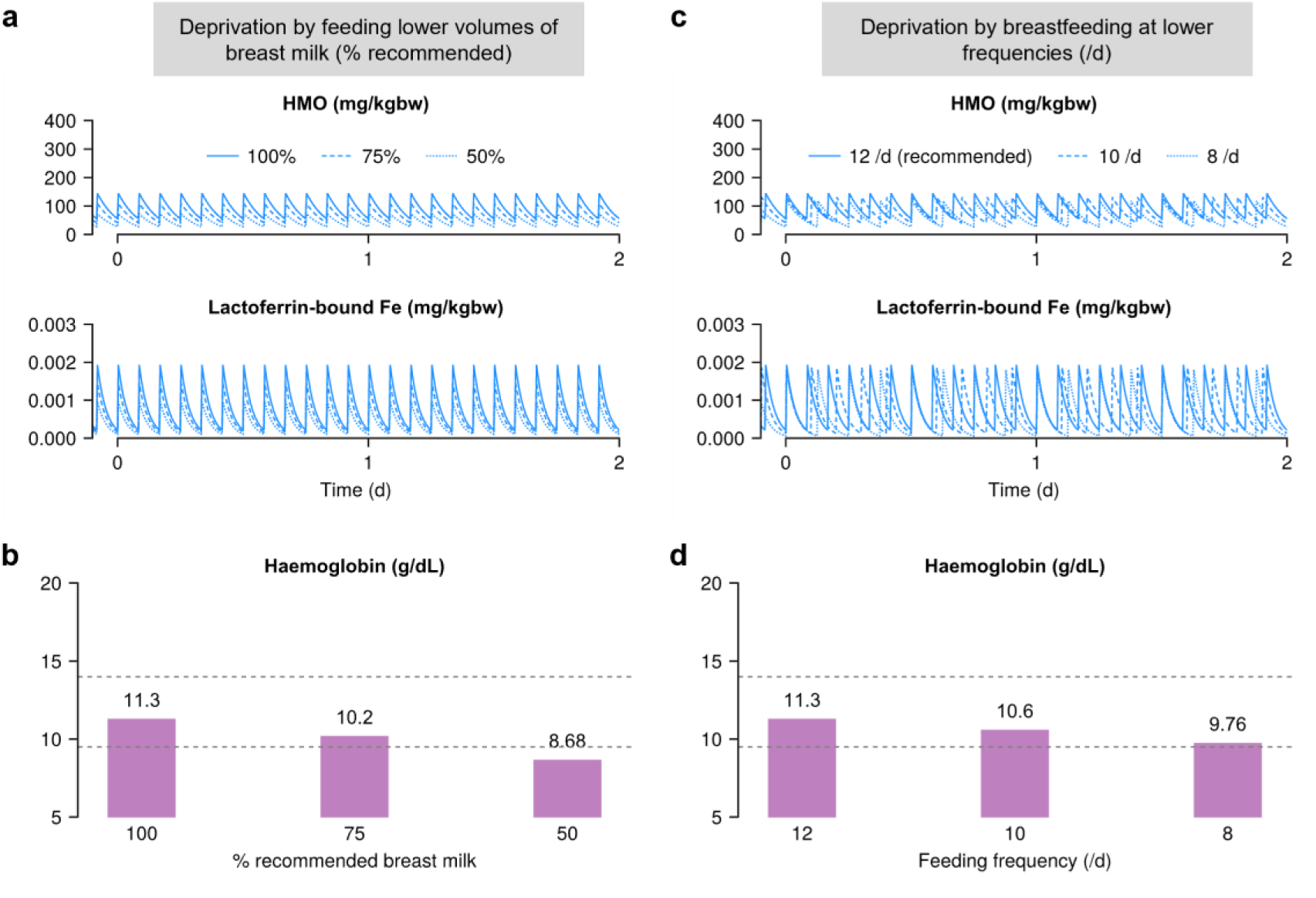
Model predictions of iron homeostasis and deficiency for a representative 4-month-old child. Deprivation of iron, either due to lower daily breast milk intake (**(a)**–**(b)**) or feeding at lower frequencies (**(c)**–**(d)**), simulated for a 4-month infant. HMO and lactoferrin-bound Fe from breast milk when 100% (solid), 75% (dash), and 50% (dot) of the recommended quantities were fed per feed **(a)** and the associated haemoglobin levels **(b)**. **(c)**–**(d)** The same quantities as in **(a)**–**(b)** but for different feeding frequencies: 12 /d (solid), 10 /d (dash), and 8 /d (dot). The horizontal dashed lines indicate the normal ranges (Table S3).

When we performed similar calculations with representative 18-(Fig. S4) and 24-month-old (Fig. S5) children, accounting for their distinct dietary sources, our model recapitulated iron homeostasis with normal diet as well as ID following dietary deprivation (Figs. S4 and S5). Sensitivity analysis showed that our predictions of haemoglobin levels were robust to variations in parameter values (Fig. S6).

### Alleviation of iron deficiency with iron-fortified nutritional solutions

We next examined whether the above ID could be addressed using iron-fortified nutritional solutions. We considered nutritional solutions specifically designed for children of the three age groups (Table S4) and simulated their administration under the feed volume (Fig. 3a, Fig. S7) or frequency (Fig. 3b, Fig. S8) deprivation conditions described above. Our model predicted that with continued administration haemoglobin levels were restored to the normal range when the deprivation was modest (Figs. 3a and b). For instance, in a 4-month infant, the model predicted that the nutritional solution (Table S4) would address the deficit when the breast milk volume was reduced to 75% of the normal volume but not when reduced to 50%.

**Fig. 3.**
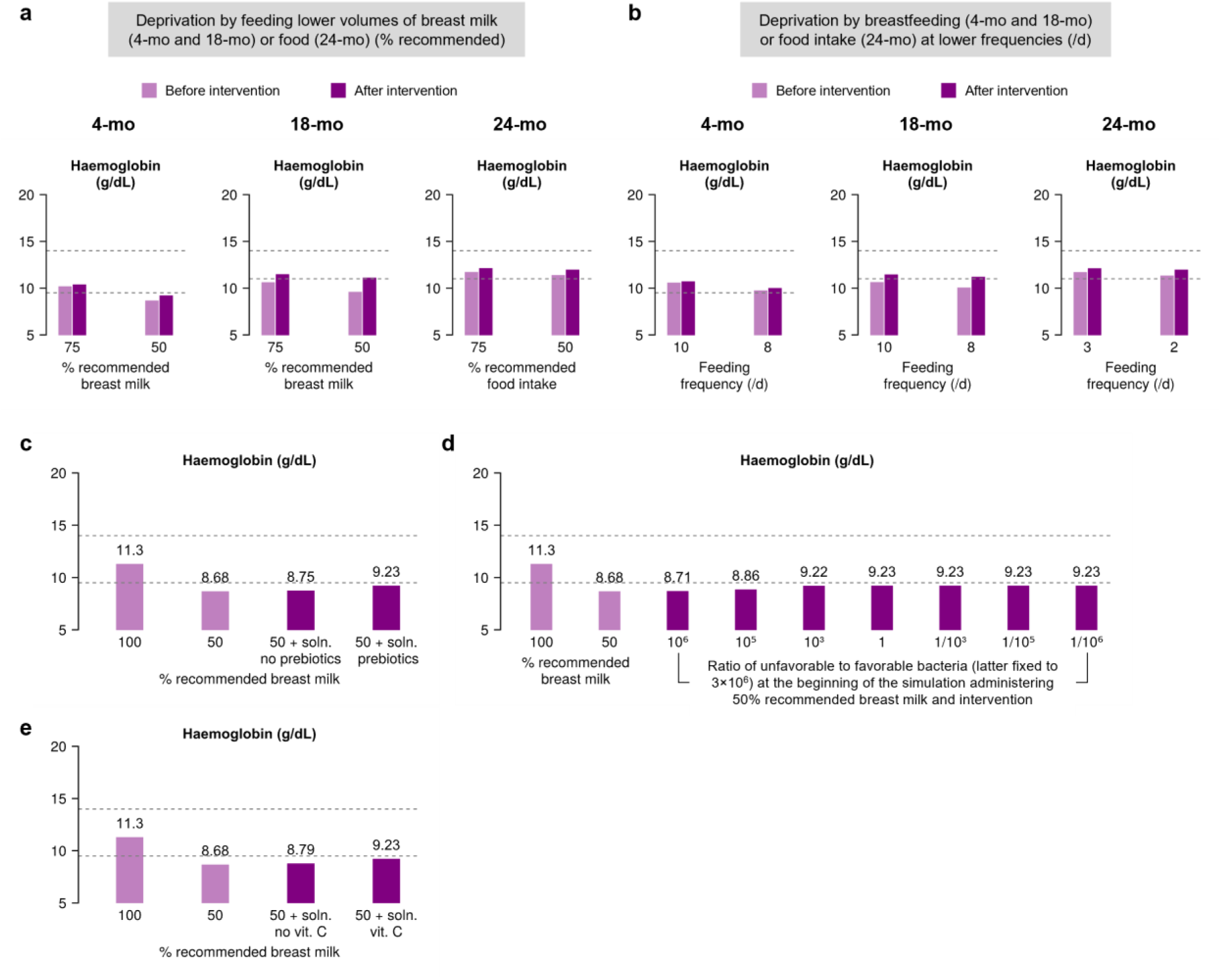
Nutritional solution and the role of its ingredients in addressing iron deficiency. Children deprived of iron in their feeds (4-mo: only breast milk, 18-mo: breast milk and food, and 24-mo: only food), either due to lower levels of daily intake **(a)** or feeding at lower frequencies **(b)**, were provided with iron-fortified nutritional solutions (Table S4). The deprivation conditions tested were the same as in Fig. 2. The effect of **(c)** prebiotics, **(d)** ratio of unfavorable and favorable gut bacteria, and **(e)** vitamin C on iron absorption from iron-fortified nutritional solutions in a typical 4-month-old infant. The horizontal dashed lines indicate the normal ranges (Table S3).

To understand the restoration better, we assessed the roles of the major ingredients in the nutritional solution known to influence iron absorption: prebiotics (FOS and inulin) and vitamin C. In the absence of prebiotics, the restoration of haemoglobin levels could be inadequate, reaching ∼8.8 g/dL instead of the ∼9.2 g/dL with prebiotics in a typical 4-month-old infant receiving 50% of the standard breast milk volume (Fig. 3c). Non-absorbed iron from the supplement can preferentially aid the growth of unfavourable bacteria, which may adversely affect favourable bacteria, compromising gut pH and hence iron absorption.

Prebiotics restore the balance of gut bacteria and facilitate iron absorption. Expectedly, the restoration was sensitive to the relative abundance of unfavourable bacteria in the gut. An increase in their abundance beyond an unfavourable:favourable bacterial ratio of 10^3^:1 hampered iron absorption substantially, further highlighting the role of the gut microbial composition, and thus the importance of prebiotics in the iron-fortified nutritional solutions (Fig. 3d). Similarly, when the nutritional solution lacked vitamin C, haemoglobin levels were not adequately restored (Fig. 3e). These results suggest that both prebiotics and vitamin C are important ingredients in nutritional solutions.

Together, these model predictions helped understand iron homeostasis, ID and iron-fortified nutritional solutions at the individual infant level. We examined next these effects at the population level.

### Model captures clinical trial data

We asked whether our model could recapitulate observations of ID and the influence of iron-fortified nutritional solutions in a clinical trial setting. We considered the Benefits and Risks of Iron Interventions in Children (BRISC) study^31^, a large clinical trial in Bangladesh. In the study, 3300 8-month-old children from rural Bangladesh who were either mildly anaemic or non-anaemic (haemoglobin levels at baseline: 11.0±1.0 g/dL) were randomized into three groups of about 1100 children each. The first group was given a daily dose of iron syrup containing 12.5 mg of elemental iron as Fe^2+^ (ferrous sulphate) salt for 3 months. The second group was given a daily dose of multiple micronutrient powder (MNP), which contained 12.5 mg of elemental iron as Fe^2+^ (ferrous fumarate) salt along with other nutrients like folic acid, zinc, and vitamins (including vitamin A and vitamin C), for the same duration. The third group received a placebo which had no iron. Haemoglobin levels at baseline and at the end of the intervention were measured.

We simulated the BRISC trial as follows. We first generated a large virtual cohort of 8-month-old children by accounting for inter-individual variations in body weight, volume of breast milk consumed, concentration of iron in breast milk, and the ratio of favourable to unfavourable bacteria in the gut microbiota (Methods, Fig. S9). We applied our model to each infant and predicted haemoglobin levels. From these virtual children, we selected 3,300 children with haemoglobin levels in the range 9 – 13 g/dL and forming a normal distribution with mean ∼11.0 g/dL and standard deviation 1.0 g/dL (Table S5, Fig. 4a). This yielded a baseline virtual population that matched the study. These virtual children were allotted to one of the three groups in roughly equal numbers so that the baseline haemoglobin levels and the fraction of children who were anaemic (haemoglobin levels <11 g/dL) in each group were consistent with the baseline data in the trial (Fig. 4a). We then applied our model to predict the outcomes of intervention. We introduced 12.5 mg of Fe^2+^ per day to the first two groups, mimicking the interventions for groups 1 and 2 in the study. The third group received no iron as part of the intervention.

**Fig. 4.**
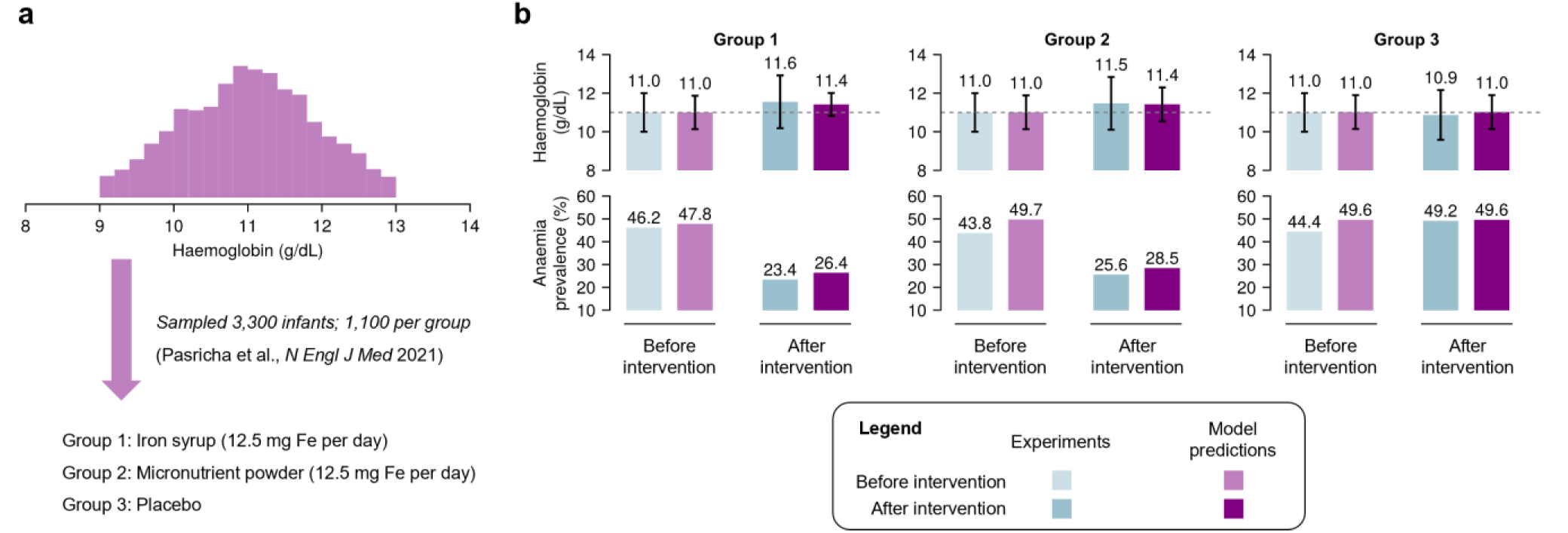
Model recapitulates clinical trial observations. **(a)** Distribution of haemoglobin levels in a virtual population of 3,300 8-month-old children (Methods, Fig. S9). The children are randomly allotted to one of the three intervention groups in equal numbers. **(b)** Mean haemoglobin levels (top row) and percentage anaemic individuals (bottom row) measured before and after the intervention in the three groups (blue) and compared to the model predictions (purple). Numbers above the bars indicate their height. Error bars are standard deviations. The horizontal dashed line indicates the lower limit of the normal range (Table S3).

In the trial, about half of the children in the groups were anaemic before intervention (46.2% in group 1, 43.8% in group 2; and 44.4% in group 3; Fig. 4b). The groups all had median haemoglobin levels of ∼11 g/dL (Fig. 4b). At the end of 3 months, groups 1 and 2 showed an increase in haemoglobin levels of 0.55±0.93 g/dL and 0.47±0.93 g/dL, respectively (Table S5, Fig. 4b). Furthermore, the prevalence of anaemia dropped to ∼23.4% in group 1 and 25.6% in group 2, demonstrating recovery of nearly half the anaemic individuals (Fig. 4b). In excellent agreement, our model predicted, without any adjustable parameters, an increase in the haemoglobin level of 0.42±1.21 g/dL and 0.42±1.25 g/dL in groups 1 and 2, respectively. Furthermore, the percentage of anaemic children decreased from 47.8% to 26.4% in group 1 and 49.7% to 28.5% in group 2. Consistent with the trial data, no significant change was observed in the placebo arm. This close agreement of our model predictions with the trial data gave us confidence in our model. We applied it next to predict the effect of nutritional solutions on wider, real-world populations.

### Benefits of iron-fortified nutritional solutions in real world settings: Application to India

Nutritional solutions can be administered without prescription to mildly anaemic children to reduce iron deficiency and alleviate associated anaemia; moderate and severe anaemia require prescription medication (see Table S2 for these categories). Nutritional solutions in the form of formula feeds are widely available. We therefore asked whether administering such solutions to mildly anaemic individuals would be beneficial. To answer this question, we focused on Indian children, for which the parameters used to describe the BRISC trial are expected to hold, given their similar ethnic, demographic, and socio-economic backgrounds (Fig. 4a, Table S2). Following the procedure above, we developed virtual populations (Methods), with 10,000 individuals each for the three age groups we studied above (Fig. S9). We excluded individuals with haemoglobin levels either too low (<7 g/dL) or too high (>17 g/dL) (Fig. S10). We ensured that the proportion of children in our virtual population with normal haemoglobin levels (32.9%) and anaemia (mild: 29%, moderate: 36%, and severe: 2.1%) matched the recent National Family Health Survey-5 (NFHS-5)^3^ estimates for Indian children.

Three different iron-fortified nutritional solutions are recommended for children in the three age groups (Table S4). Applying our model with the ingredients in the respective solutions administered at the recommended dosages, we found remarkably that a substantial number of mildly anaemic children had their haemoglobin levels eventually restored to normalcy (Fig. 5, Table S5). In the 4-month-old group, the mean haemoglobin level in the mildly anaemic infants rose from 9.27 g/dL to 9.64 g/dL, the latter in the normal range for this age group. Indeed, about 85% of the mildly anaemic children had their haemoglobin levels restored to the normal range in our predictions. For the 18-month-old group, the mean rose from 10.5 g/dL to 11.4 g/dL, the latter well into the normal range for this age group.

**Fig. 5.**
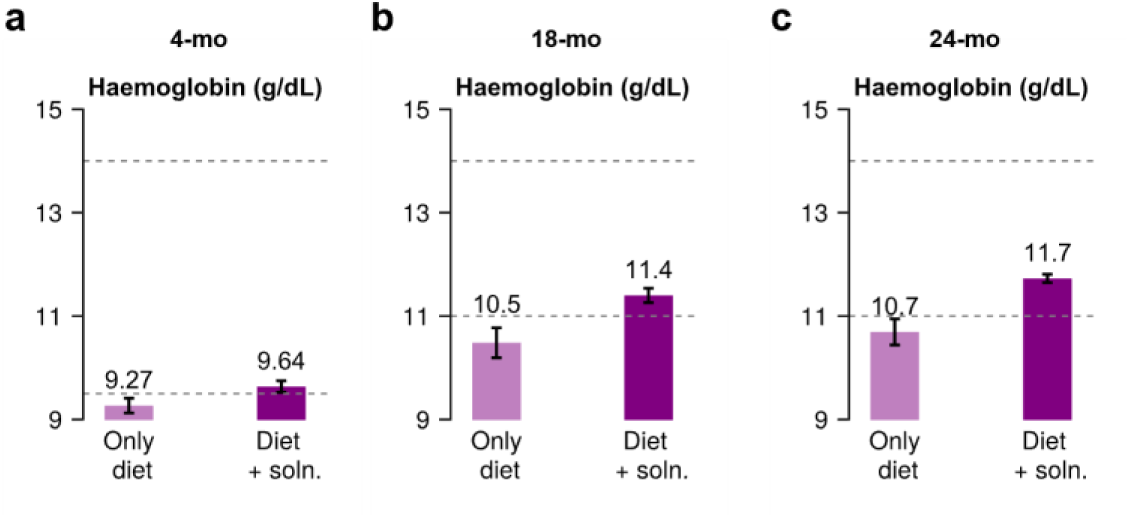
Predicted effect of nutritional solutions on children populations. Mean haemoglobin levels in children before (lighter bar) and after (darker bar) intervention with nutritional solutions in 4-mo **(a)**, 18-mo **(b)**, and 24-mo **(c)** age groups. Numbers above the bars indicate their heights. Error bars are standard deviations. The horizontal dashed lines indicate the normal ranges (Table S3).

Similarly, for the 24-month-old group, the mean rose from 10.7 g/dL to 11.7 g/dL, the latter again in the normal range. For the latter two groups, remarkably, over 95% of the mildly anaemic children had their haemoglobin levels restored to the normal range. In healthy infants, haemoglobin levels were predicted to rise only modestly, remaining in the normal range (Fig. S11). In sum, our study predicts that the usage of iron-fortified nutritional solutions may greatly benefit in alleviating ID in mildly anaemic children, a public health intervention that may be widely and readily deployed given their over-the-counter availability. It also offers a framework to test alternative nutritional solutions that may be designed to achieve further improvements in recovery rates.

## Discussion

Studies over the years have identified key molecular, cellular, and systemic players involved in iron homeostasis^5,12^. The resulting insights have helped develop interventions to address ID that may result from inadequate dietary supply and/or the inability of these regulatory mechanisms to maintain homeostasis. For instance, introducing prebiotics to aid iron absorption in the gut draws upon the knowledge of the role of the gut microbiome in regulating gut pH, which in turn influences the functioning of the metal-absorbing proteins expressed on enterocytes^25^. Yet, the interconnected nature of the many regulatory processes involved, across cellular and systemic levels, demands a systems-level view of iron homeostasis that synthesizes these processes and helps assess the influence of interventions. Our study fills this knowledge gap. It presents, to our knowledge, the first computational model that offers a comprehensive, quantitative framework of paediatric iron homeostasis and helps evaluate the impact of interventions.

We built the model to recapitulate paediatric anaemia and the impact of interventions in the Indian subcontinent. Remarkably, the model captured quantitatively, without adjustable parameters, the effects of deploying nutritional interventions observed in a large clinical trial in Bangladesh, demonstrating its accuracy, robustness, and utility. Furthermore, it predicts that available iron-fortified nutritional solutions, including over-the-counter products designed to foster iron absorption, may greatly benefit mildly anaemic children. Specifically, in a paediatric population under 2 years of age reflecting the latest national survey in India, over 85% of the mildly anaemic children were predicted to have their haemoglobin levels restored with age-appropriate over-the-counter solutions, thus suggesting a potentially important public health intervention.

Our study opens avenues for future research. We considered ID arising from deficient supply of iron. Our model could be extended to other causes of ID, such as those arising from poor dietary absorption of iron due to gut dysbiosis or chronic inflammation^5^.

Interventions would then have to consider ingredients beyond standard nutritional solutions. For instance, a higher prevalence of pathogenic bacteria in the gut may worsen anaemia if iron levels in the nutritional solutions are not carefully regulated because excess iron is consumed by pathogenic gut bacteria, compromising the growth of commensals and hence iron absorption^32^. Thus, introducing pre- and/or pro-biotics together with a tailored proportion of iron may be optimal in such scenarios. Our model offers an avenue to evaluate such interventions and devise optimal ones. In the same way, while our model has been developed for paediatric settings, we anticipate extending it to adults. An important group is women, who tend to have a high incidence of iron deficiency anaemia^1^.

We recognize limitations of our study. First, we chose model parameter values from a careful study of the literature. Yet, several parameter values remain unknown and were set to values that ensured recapitulation of homeostatic markers in healthy children. We tested our model predictions for their sensitivity to these parameter values and found them to be robust. Yet, we cannot rule out alternative parameter combinations that may yield similar results. Future studies may assess this possibility once estimates of the unknown parameters become available. Second, we realize that some biological processes in iron homeostasis are yet to be fully understood^12^. For instance, the mechanism of absorption of heme-iron by enterocytes is yet unknown. Here, for simplicity, we have ignored such processes. We anticipate the effect of the resulting approximations to be minimal given that our model captured clinical data accurately. Third, although our model is complex, it is minimal on many fronts. We chose simple empirical approximations to capture many complex underlying processes, so that the model parameter space remained tractable. For instance, iron absorption happens at different rates in different sections of the gut lumen^33^. Here, we modelled the gut as a single compartment with an iron absorption rate that is the average of the rates in different sections. Similarly, children are born with iron stores that typically meet the physiological iron requirements during their early growth^34^. Here, we ignored the contribution from iron stores because the stores are depleted by 4–6 months of age^35^. Our calculations were performed for children of age 4 months and above. Future studies may account for the role of stores to characterize iron metabolism in younger infants. Fourth, the empirical link we chose between serum transferrin-bound iron levels and steady state haemoglobin levels in healthy individuals may help accurately predict steady states but at the expense of dynamics (Methods), which is an acceptable trade-off given our goal of predicting the final haemoglobin levels realized under different settings including with iron-fortified nutritional solutions. Future advances may replace the empirical forms with mechanistic descriptions, enabling the application of our model to devise not only better nutritional solutions but also optimize durations of their use. Finally, we recognize that our model did not consider potential long-term implications of supplementation especially when administered to children whose haemoglobin levels are in/have reached normal levels; supplementation in children with already high levels of haemoglobin levels may trigger adverse developmental outcomes^10^.

In summary, our study presents a comprehensive mathematical model of iron homeostasis in children that helps understand the interplay of the various regulatory mechanisms in maintaining iron homeostasis and predicts the effect of supplementation via iron-fortified nutritional solutions in alleviating ID at the individual- and population-levels.

## Methods

### Mathematical model

We construct the following equations to describe the processes involved in iron homeostasis and supplementation in children. The equations are grouped by the main compartments they apply to, namely, gut lumen, enterocytes, circulation, and the nutritional solution.

*Gut lumen*

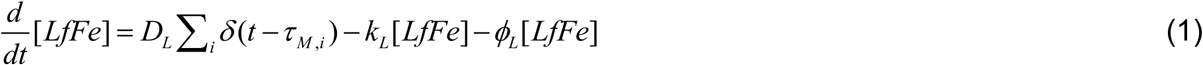

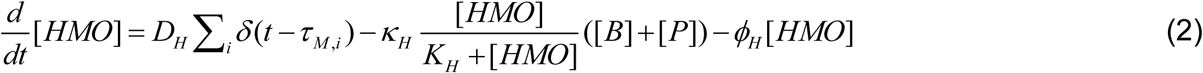

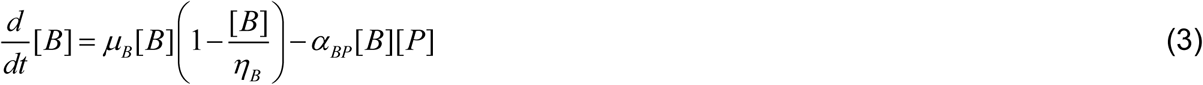

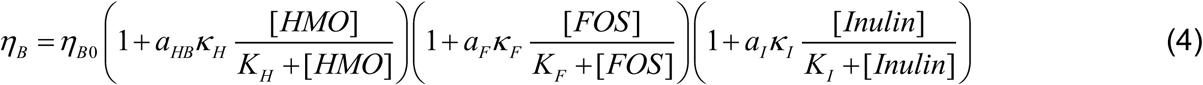

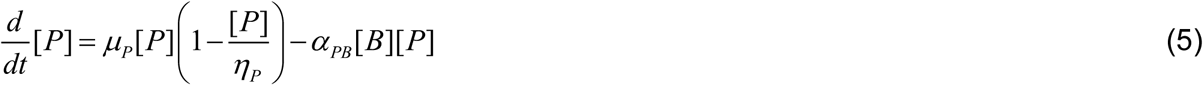

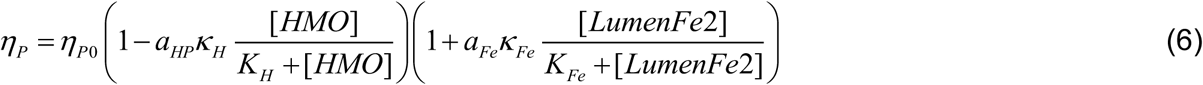

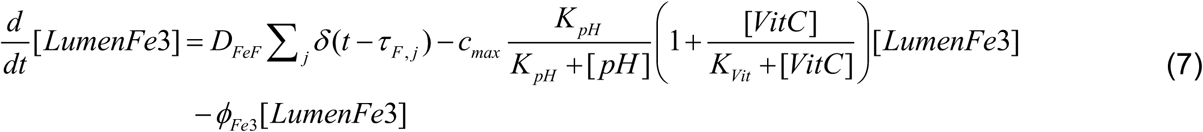

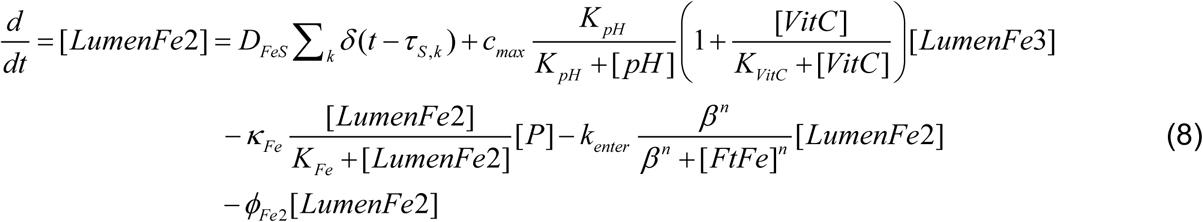

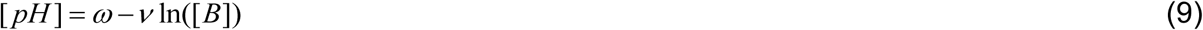

*Enterocytes*

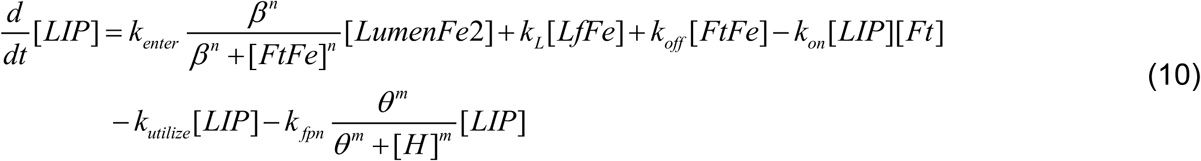

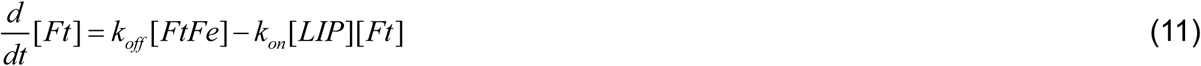

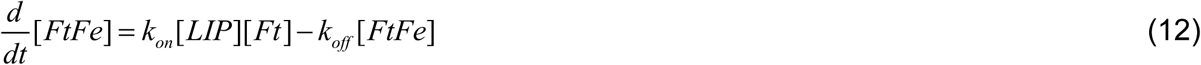

*Circulation*

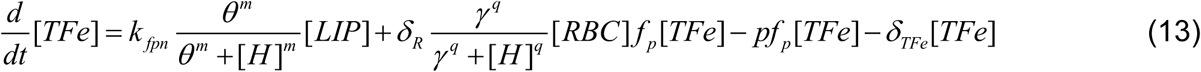

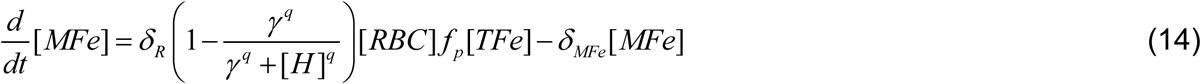

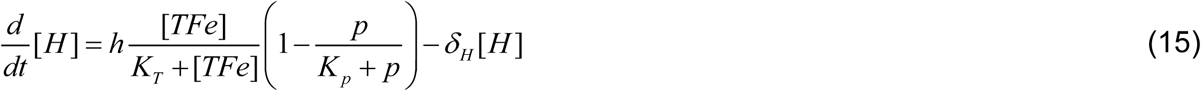

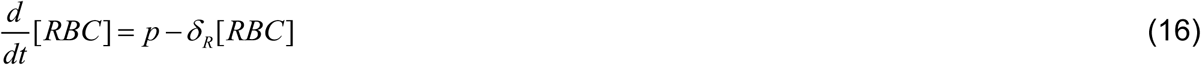

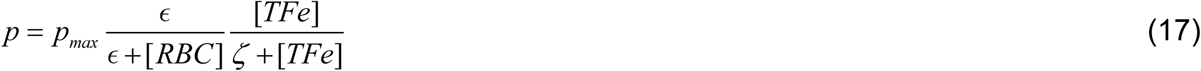

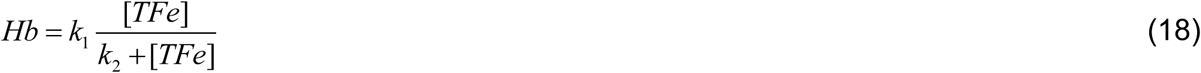

*Nutritional solution*

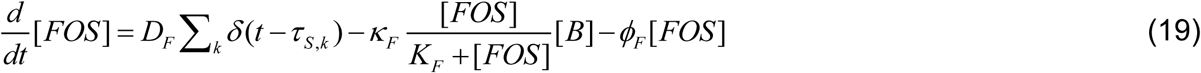

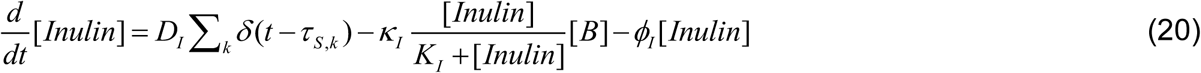

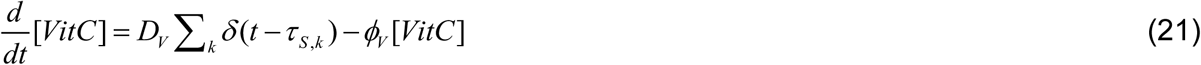

We now describe the equations term-by-term.

Breast milk introduces lactoferrin-bound Fe into the gut lumen. Feeding is assumed to occur at time points denoted *τ _M_* _,*i*_, at which times an amount *D_L_* of lactoferrin-bound iron, denoted [*LfFe*], is added to the gut lumen (Eq. 1). The Dirac delta function ensures the introduction at these times, as *δ* (*x* ≠ 0) = 0 and 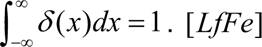 is absorbed by enterocytes with the per capita rate *k_L_* and cleared from the lumen with the per capita rate *ϕ_L_*.

Breast milk also introduces human milk oligosaccharides, denoted [*HMO*], into the gut lumen. We let an amount *D_H_* be introduced at each dose, along with [*LfFe*] (Eq. 2). [*HMO*] is consumed by gut bacteria (favourable, [*B*], and unfavourable, [*P*]) at the maximal per capita rate *κ _H_* and half-maximal constant *K_H_*, and is cleared from the lumen with the per capita rate *ϕ_H_*.

We group the gut microbial species into favourable (commensal) and unfavourable (pathogenic) categories, for simplicity. The favourable bacteria, [*B*], are assumed to grow following a logistic term with per capita growth rate *μ_B_* and carrying capacity *η_B_* (Eq. 3). Their growth is suppressed by competition with pathogenic bacteria, [*P*], with the interaction rate constant *α_BP_*, following standard models of bacterial competition^36,37^. We let the carrying capacity *η_B_* be a function of the available resources in the gut. Thus, the carrying capacity increases from its basal level, *η_B_*_0_, with the level of [*HMO*] consumed, with *^a^_HB_* being the proportionality constant (Eq. 4). Similarly, [*FOS*] and [*Inulin*] supplied as part of nutritional solutions also contribute to this increase, with proportionality constants *a_F_* and *a_I_*, respectively (Eq. 4).

The unfavourable bacteria, [*P*], also grow following a logistic term with growth rate *μ_P_* and carrying capacity *η_P_*, and compete with the favourable bacteria with the interaction strength *α_PB_* (Eq. 5). Further, the carrying capacity of these bacteria decreases from the basal rate *η_P_*_0_ with the rate of [*HMO*] consumption, with the proportionality constant *a_HP_* (Eq. 6). The carrying capacity increases also with the amount of Fe^2+^ in the lumen, [*LumenFe*2], consumed by the bacteria, with *a_Fe_* being the proportionality constant (Eq. 6).

We next examine iron coming into the gut from other food sources. We consider food intake at times *τ _F_*, *_j_*, each time introducing an amount *D_FeF_* of iron in the form of Fe^3+^ into the lumen, denoted [*LumenFe*3] (Eq. 7). Similarly, nutritional solutions, administered at times *τ _S_*_,*k*_, introduce the amount *D_FeS_* of Fe^2+^ to [*LumenFe*2]. DcytB converts Fe^3+^ to Fe^2+^ in a gut pH dependent manner with the maximum per capita rate *^c^_max_* and the half-maximal constant *K _pH_*. This rate is enhanced by vitamin C, [*VitC*], again in a saturable manner with half-maximal constant *K_Vit_*. [*LumenFe*3] is cleared from the gut lumen with the per capita clearance rate *ϕ_Fe_*_3_.

[*LumenFe*2] is taken up by unfavourable bacteria at the maximal per capita rate *κ_Fe_* and half-maximal constant *^K^_Fe_* (Eq. 8). It enters enterocytes at the per capita rate *k_enter_*, which is negatively regulated by ferritin-bound iron stores in the enterocytes, [*FtFe*], with the half-maximal constant *β* and the Hill coefficient *n*. It is cleared from the gut lumen with the per capita rate *ϕ_Fe_*_2_.

Finally, we recognize that the gut pH, denoted [*pH*], is lowered by favourable gut bacteria from its nominal value *ω* to a value dependent on the logarithmic abundance of the bacteria, [*B*], with the proportionality constant *ν* (Eq. 9).

The above equations describe the processes in the gut lumen. We describe next the processes in enterocytes. The absorbed Fe^2+^ from the lumen enters the labile iron pool, [*LIP*], in the enterocytes (Eq. 10). This includes lactoferrin-bound Fe as well as that absorbed by DMT1. Iron from the labile pool can be utilized for cellular functions with the per capita rate *k_utilize_*, can be stored reversibly in ferritin cages with the binding and unbinding rate constants *^k^_on_* and *k_off_*, respectively, and be exported via ferroportin to circulation with the per capita rate *k _fpn_*. The latter rate is negatively regulated by circulating hepcidin levels, [*H*], with the half-maximal constant *θ* and Hill coefficient *m* (Eq. 10). The reversible binding of iron to ferritin cages dictates the evolution of free ferritin, [*Ft*], and iron bound to ferritin, [*FtFe*] (Eqs. 11 and 12, respectively).

We next describe the processes in circulation. The export of iron from enterocytes contributes to the transferrin bound iron in circulation, [*TFe*] (Eq. 13). A major source of [*TFe*] is recycling of iron from senescent RBCs via macrophages. We let this rate be proportional to the death rate of RBCs, *δ _R_*[*RBC*], and the amount of iron contained in each RBC, the latter determined as *f _p_* [*TFe*], the amount of transferrin bound iron taken up by each RBC during erythropoiesis. We recognize that RBCs have an average lifespan of 120 d^38–40^, and thus the [*TFe*] in the latter term should be the value of [*TFe*] 120 d in the past.

Modelling the age-structured RBC population balance is required for predicting the dynamics of haemoglobin levels changes^41^. Here, our interest is in predicting the long-term or steady-state level of haemoglobin, for which employing the current value of [*TFe*] works well. The export of iron from macrophages is also regulated by hepcidin, with the half-maximal constant *γ* and Hill coefficient *q*. [*TFe*] is lost from circulation predominantly due to uptake by RBCs during erythropoiesis. We let this rate be proportional to the production rate of RBCs, *p*, and the amount of transferrin bound iron taken up by each RBC, *f _p_* [*TFe*]. We let [*TFe*] be cleared from circulation with the per capita rate *δ_TFe_*, which includes its transport to all sites other than the bone marrow.

The iron in macrophages not exported to circulation is stored in the macrophages, [*MFe*], which in turn can be lost due to macrophage turnover with the per capita rate *δ_MFe_* (Eq. 14). Hepcidin production is assumed to increase with [*TFe*] with the maximal rate ℎ and half-maximal constant *K_T_* (Eq. 15). Further, we account for the suppression of hepcidin production associated with erythropoiesis via the hormone erythroferrone as a saturable function of the RBC production rate *p* and the half-maximal constant *K_p_*. We let hepcidin be cleared with the per capita rate *δ_H_*.

RBC levels in circulation, [*RBC*], are determined as the balance between their production rate *p* and per capita death rate *δ _R_* (Eq. 16). The production rate is negatively autoregulated via the hormone erythropoietin. We thus let *p* decrease from its maximum value *^p^_max_* with the [*RBC*] with the half-maximal constant ϵ(Eq. 17). Furthermore, we account for the enhanced RBC production with increasing [*TFe*] as a saturable function of [*TFe*] with half-maximal constant *ζ*.

The haemoglobin level, *Hb*, is positively associated to the level of transferrin-bound Fe in circulation, [*TFe*], at steady state. We model the association as a sigmoid function (Eq. 18). The constants *k*_1_ and *k*_2_ are chosen such that the lower and upper limits of normal [*TFe*] levels are mapped to the corresponding limits of *Hb* (Table S3).

Finally, we examine the processes associated with the ingredients in nutritional solutions. The iron in these solutions enters the lumen as described above (Eq. 7). Additionally, the solutions may introduce prebiotics, such as long-chain FOS and inulin, and vitamin C. The introduction of all these entities occurs at the times of administration, *τ _S_*_,*k*_ (Eqs. 19-21).

Each dose contributes the amounts *D_F_*, *D_I_*, and *D_V_* of long-chain FOS, inulin, and vitamin C to the gut lumen, respectively. Long-chain FOS and inulin are consumed by favourable bacteria with maximal rate constants *κ_F_* and *κ_I_*, and half-maximal constants *K_F_* and *K_I_*, respectively. The three ingredients are cleared with the per capita rates *ϕ_F_*, *ϕ_I_*, and *ϕ_V_*, respectively.

The equations (1)–(21) constitute the model of iron homeostasis in children. We solved them using parameter values listed in Table S1. Where available, the parameters were drawn from independent studies in the literature focussed on the individual processes involved. We assumed the remaining parameters values such that iron homeostasis in healthy children was recapitulated.

### Virtual populations and *in silico* simulations of iron-fortified nutritional solutions

We created virtual populations of 10,000 children in each of the three age groups: 4-month-olds, 18-month-olds, and 24-month-olds. For each group, we accounted for variations in body weight^42^, breast feed amounts^28^ and its iron content^30^, iron intake from other dietary sources^43^, as well as the levels of favourable and unfavourable gut bacteria. Values for these variables associated with each individual were sampled randomly from underlying distributions (Table S2). We then applied our model above to each individual to predict population level variations in haemoglobin levels. We ascertained that the key quantities involved lay in clinically observed ranges (Table S3). Further, we assessed the effect of iron-fortified nutritional solutions to each group, whose ingredients are listed in Table S4. For the 4-month-old group, the iron-fortified nutritional solutions introduced 5.74 mg of iron along with 2.025 g of FOS, 4.05 g of inulin, and 54 mg of vitamin C per day. For the 18-month group, we assumed reduced breast feeding, amounting to 50% of the younger children above. Additionally, we assumed that other dietary sources offered up to 8 mg of iron per day, corresponding to the recommended dietary allowance (RDA) (Table S2). The nutritional solution, as per the recommended dosing, introduced 4.86 mg of iron and 32.4 mg of vitamin C (Table S4). Finally, the 24-month-old children, who we assumed were off breastfeeding, received all their dietary requirement of iron through other food sources. The RDA for this age group is again 8 mg (Table S2). Iron-fortified nutritional solutions introduced 3.24 mg of iron and 21.6 mg of vitamin C per day in 2 doses.

## Data Availability

This is a computational study and no new data has been generated. All calculations were performed in Julia v1.7.3. The model code and the parameter files are available at https://github.com/vembha/paediatric_iron_metabolism.

## Acknowledgments

We thank Prof. Sundar Swaminathan for comments. We thank Steven Kuijper, Britt Broersen, and their teams at Danone for critically reviewing the manuscript.

## Author contributions

SC and BV developed the computational model and led the data analysis. NMD and KVV supervised the overall research and provided critical input on model validation and interpretation. KVV, NMD, and KL conceptualized the study and contributed to data collection. AT and VPS assisted in model simulations and analysis. KP, PR, PK, VPS, and KS supported data collection. BV, SC, NMD, and KVV wrote and revised the manuscript. All authors reviewed and approved the final version of the manuscript.

## Disclaimers

The authors declare no competing interests relevant to this study.

## Ethical approval

This is a computational study using secondary data from publicly available sources. No human participants or animal subjects were involved, and therefore, ethical approval and informed consent were not required.

## Abbreviations

ID: iron deficiency, HMO: human milk oligosaccharides, DMT1: divalent metal transporter 1, DcytB: duodenal cytochrome B, LIP: labile iron pool, RBC: red blood cell, FOS: fructo-oligo saccharides, NFHS: national family health survey, RDA: recommended dietary allowance

