## Supplementary Materials for "Modelling how nutritional solutions may help alleviate paediatric iron deficiency: Application to the Indian subcontinent"

**Supplementary information details:**

Figures: 11, Tables: 5, References: 24

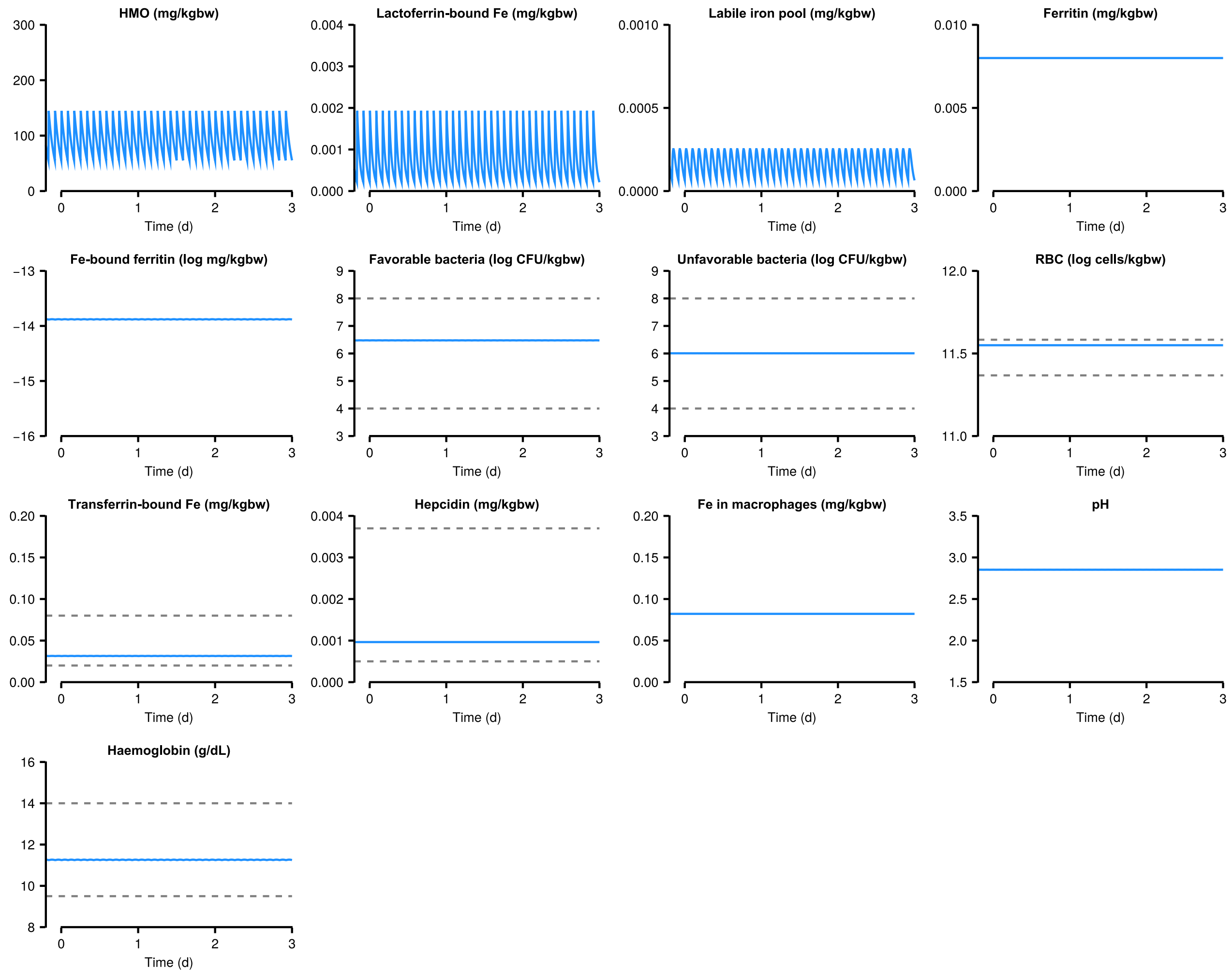


**Fig. S1. Model predictions of all variables in a 4-month-old infant under normal conditions.**

Steady state dynamics of all model variables in a 4-month-old infant receiving the recommended amount of breast milk. Model predictions are in blue and normal ranges are indicated using grey dashed lines. Normal ranges: gut bacteria ($10^4$–$10^8$ log CFU/kgbw,[1]); serum transferrin–bound Fe (0.02–0.08 mg/kgbw,[2]); hepcidin ($0.5\times10^{-3}$–$3.7\times10^{-3}$ mg/kgbw,[3]); RBCs ($23.3\times10^{10}$–$38.3\times10^{10}$ cell/kgbw,[4]); haemoglobin (10–17 g/dL,[4]).

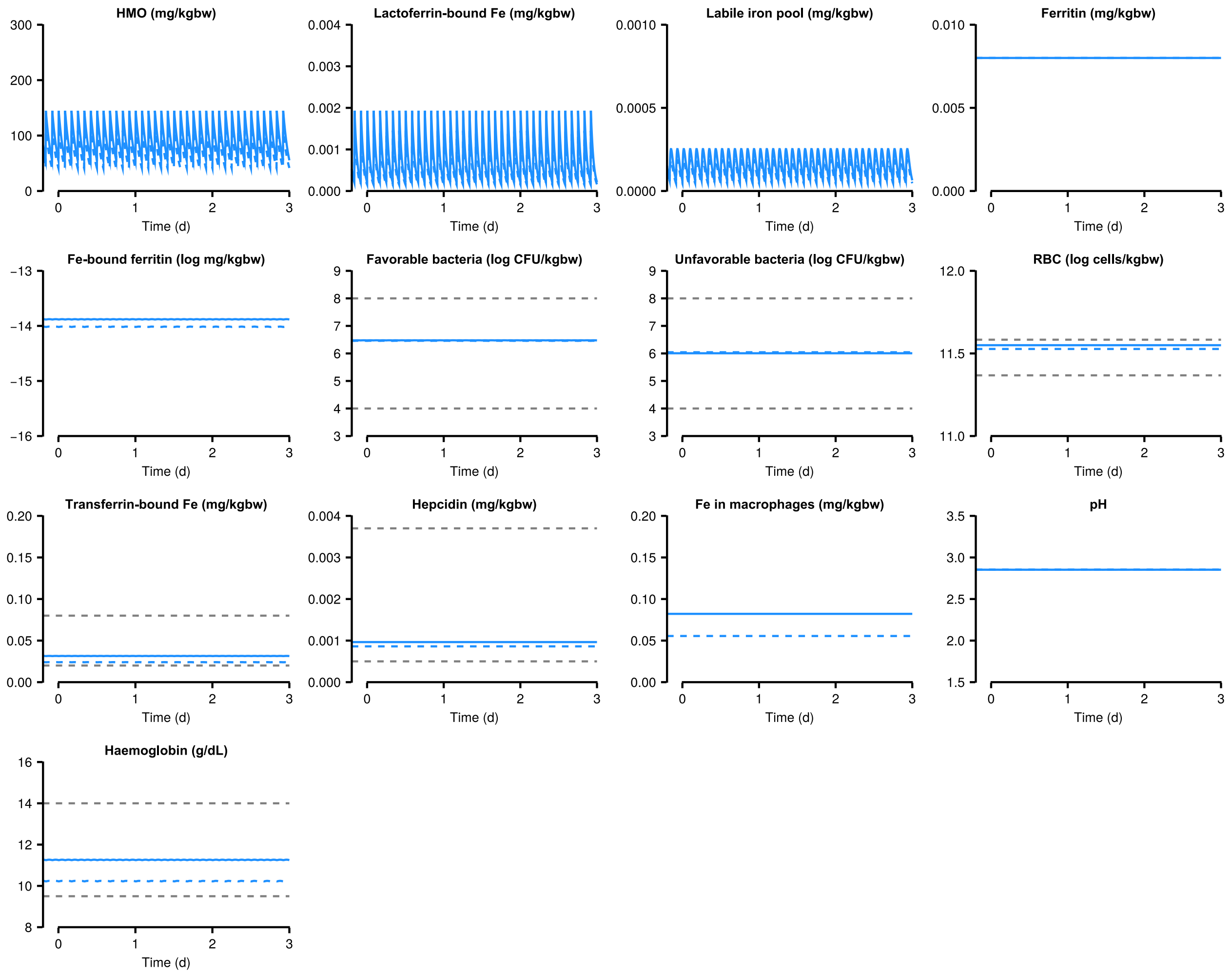


**Fig. S2. Model predictions of all variables in a 4-month-old infant receiving breast milk at deprived volume.**

Steady state dynamics of all model variables in a 4-month-old infant receiving the recommended amount of breast milk (solid) compared to that with 75% of the recommendation (dash). Model predictions are in blue and normal ranges are indicated using grey dashed lines. Normal ranges are the same as in Fig. S1.

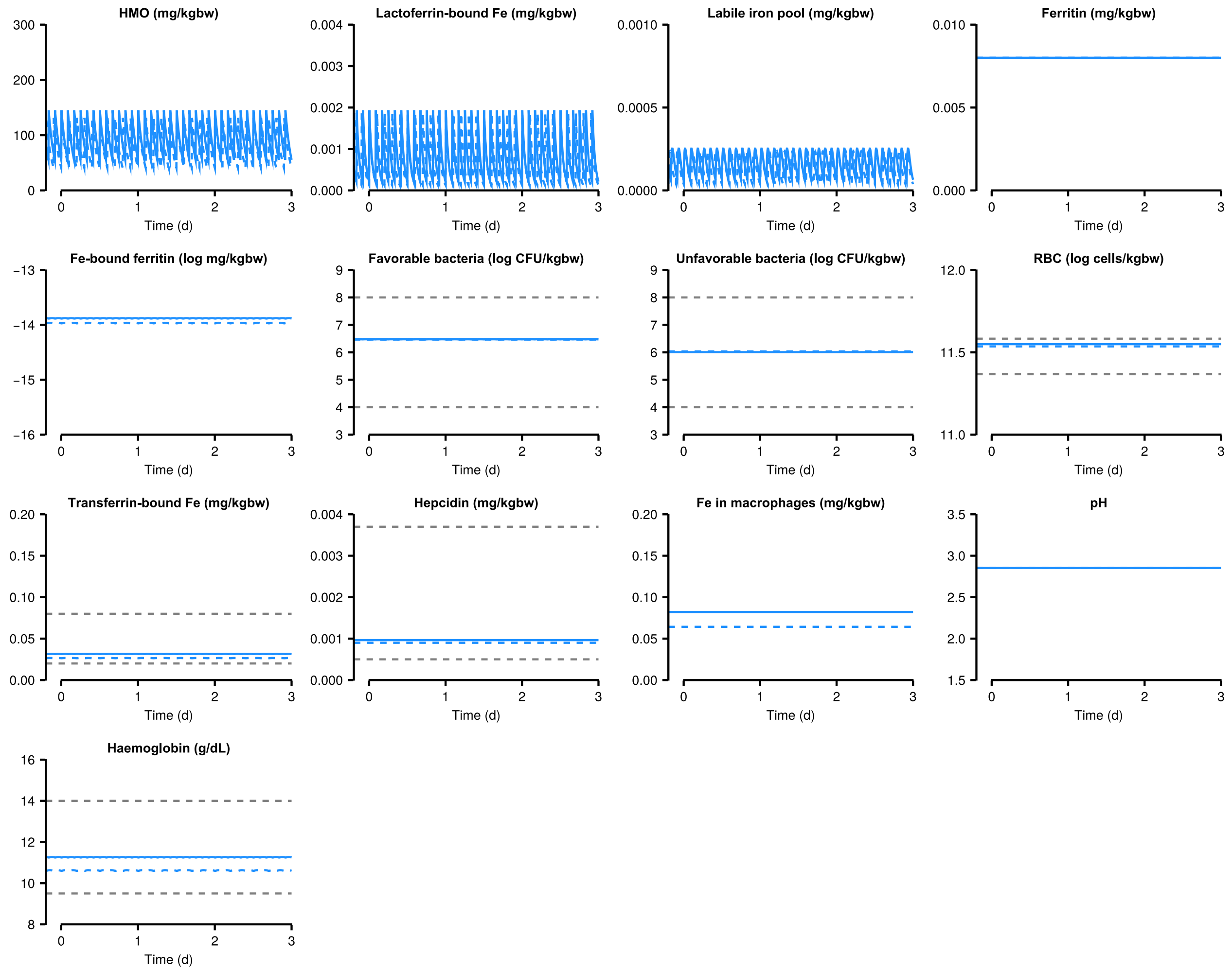


**Fig. S3. Model predictions of all variables in a 4-month-old infant receiving breast milk at deprived frequency.**

Steady state dynamics of all model variables in a 4-month-old infant receiving breast milk at the recommended frequency (solid: 12 /d) compared to that at lower frequency (dash: 10 /d). Model predictions are in blue and normal ranges are indicated using grey dashed lines. Normal ranges are the same as in Fig. S1.

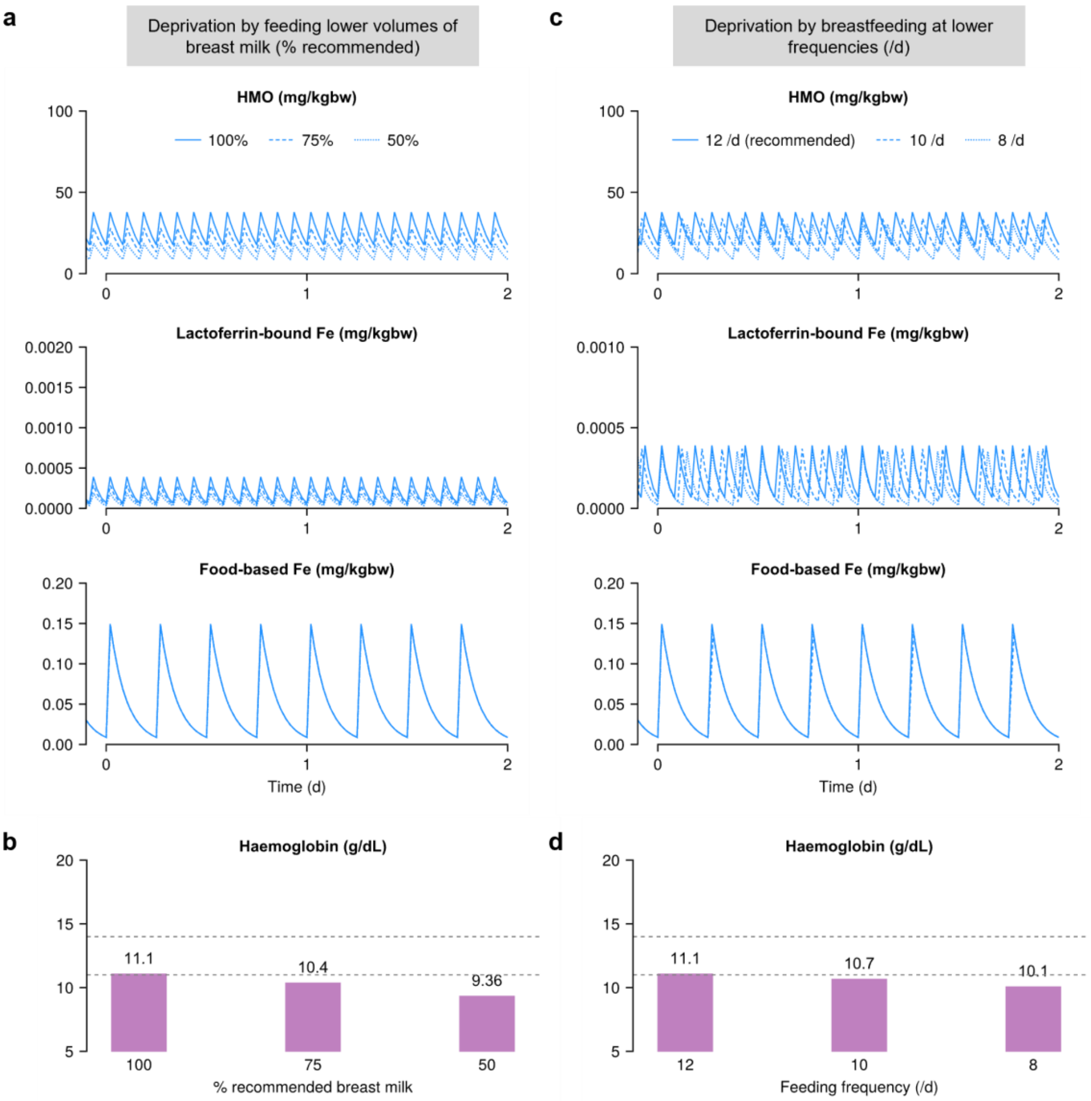


**Fig. S4. Model predictions of iron homeostasis for an 18-month-old child.**

Deprivation of iron, either due to lower daily breast milk intake (**(a)–(b)**) or feeding at lower frequencies (**(c)–(d)**), simulated for an 18-month child. HMO and lactoferrin-bound Fe from breast milk when 100% (solid), 75% (dash), and 50% (dot) of the recommended quantities were fed per feed **(a)** and the associated haemoglobin levels **(b)**. **(c)–(d)** The same quantities as in **(a)–(b)** but for different feeding frequencies: 12 /d (solid), 10 /d (dash), and 8 /d (dot). The horizontal dashed lines indicate the normal ranges (Table S3).

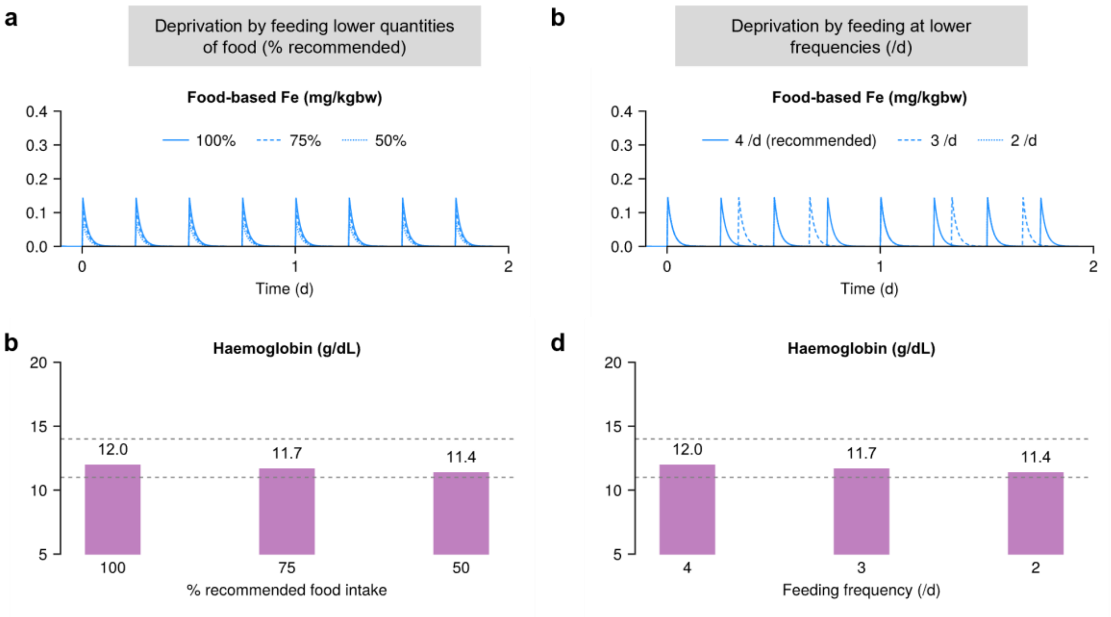


**Fig. S5. Model predictions of iron homeostasis for a 24-month-old child.**

Deprivation of iron, either due to lower daily breast milk intake (**(a)–(b)**) or feeding at lower frequencies (**(c)–(d)**), simulated for a 24-month child. Fe from food when 100% (solid), 75% (dash), and 50% (dot) of the RDA were met in four doses per day **(a)** and the associated haemoglobin levels **(b)**. **(c)–(d)** The same quantities as in **(a)–(b)** but for different feeding frequencies: 4 /d (solid), 3 /d (dash), and 2 /d (dot). The horizontal dashed lines indicate the normal ranges (Table S3).

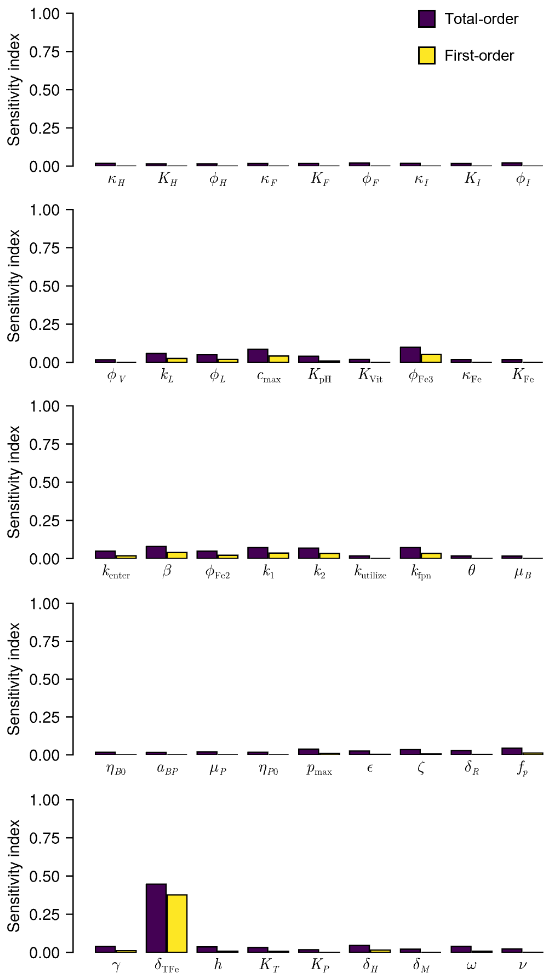


**Fig. S6. Sensitivity analysis.**

The total- and first-order sensitivity of steady state haemoglobin levels to the model parameters for 18-month age group. The eFAST routine, implemented in the GlobalSensitivity.jl package of Julia, was used to estimate the sensitivity indices using 10000 samples. Higher indices indicate greater sensitivity of heamoglobin prediction to change in the corresponding parameter values.

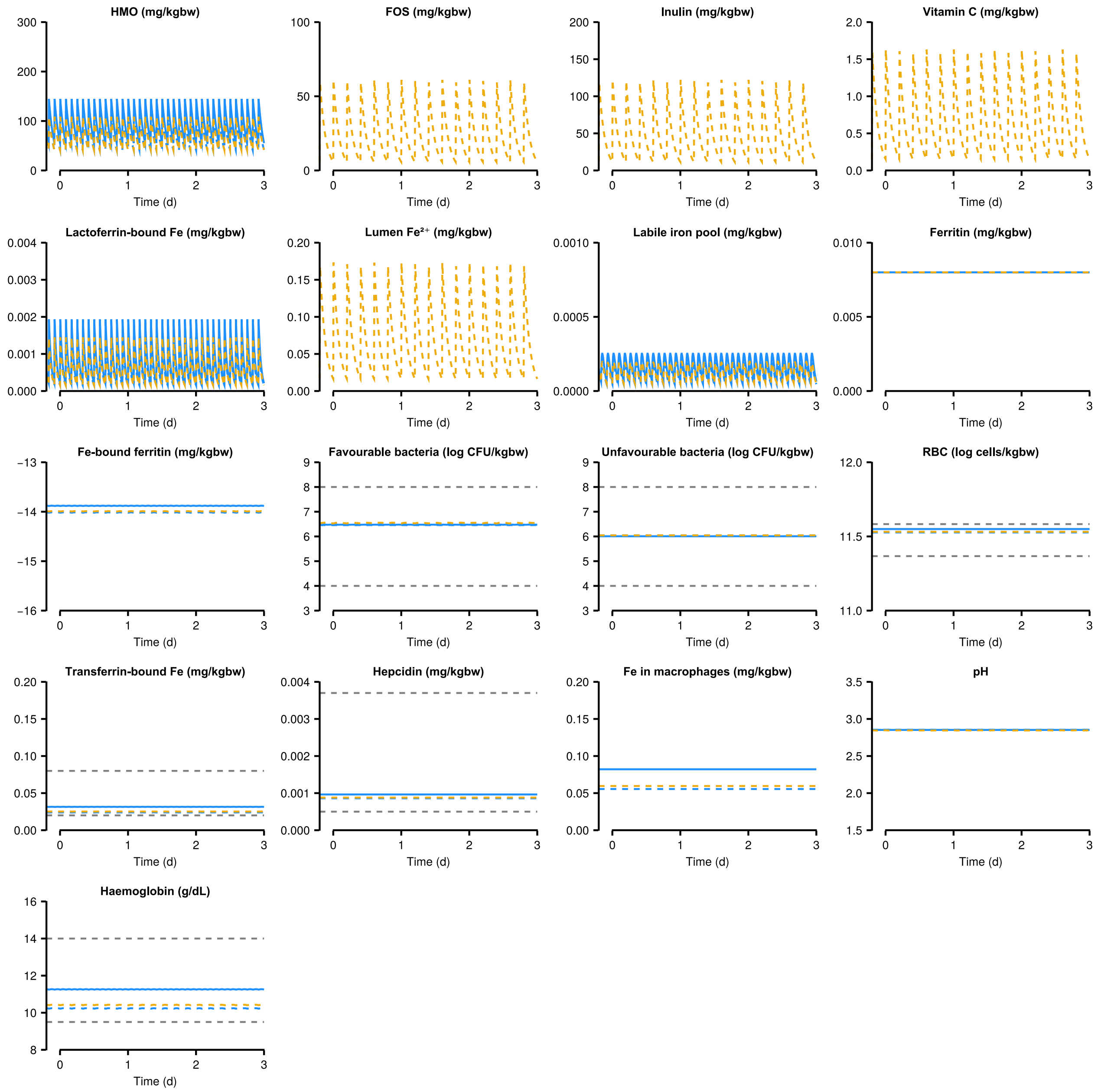


**Fig. S7. Model predictions of all variables in a 4-month-old infant receiving breast milk at deprived volume and the nutritional solution.**

Steady state dynamics of all model variables in a 4-month-old infant in Fig. S2 receiving 75% of the recommended breast milk (dash, blue) and the effect of nutritional solution (Table S4) in addressing the deprivation (dash, orange). The solid blue line indicates the predictions for the same infant receiving 100% of the recommended breast milk. Normal ranges are indicated using grey dashed lines, same as in Fig. S1.

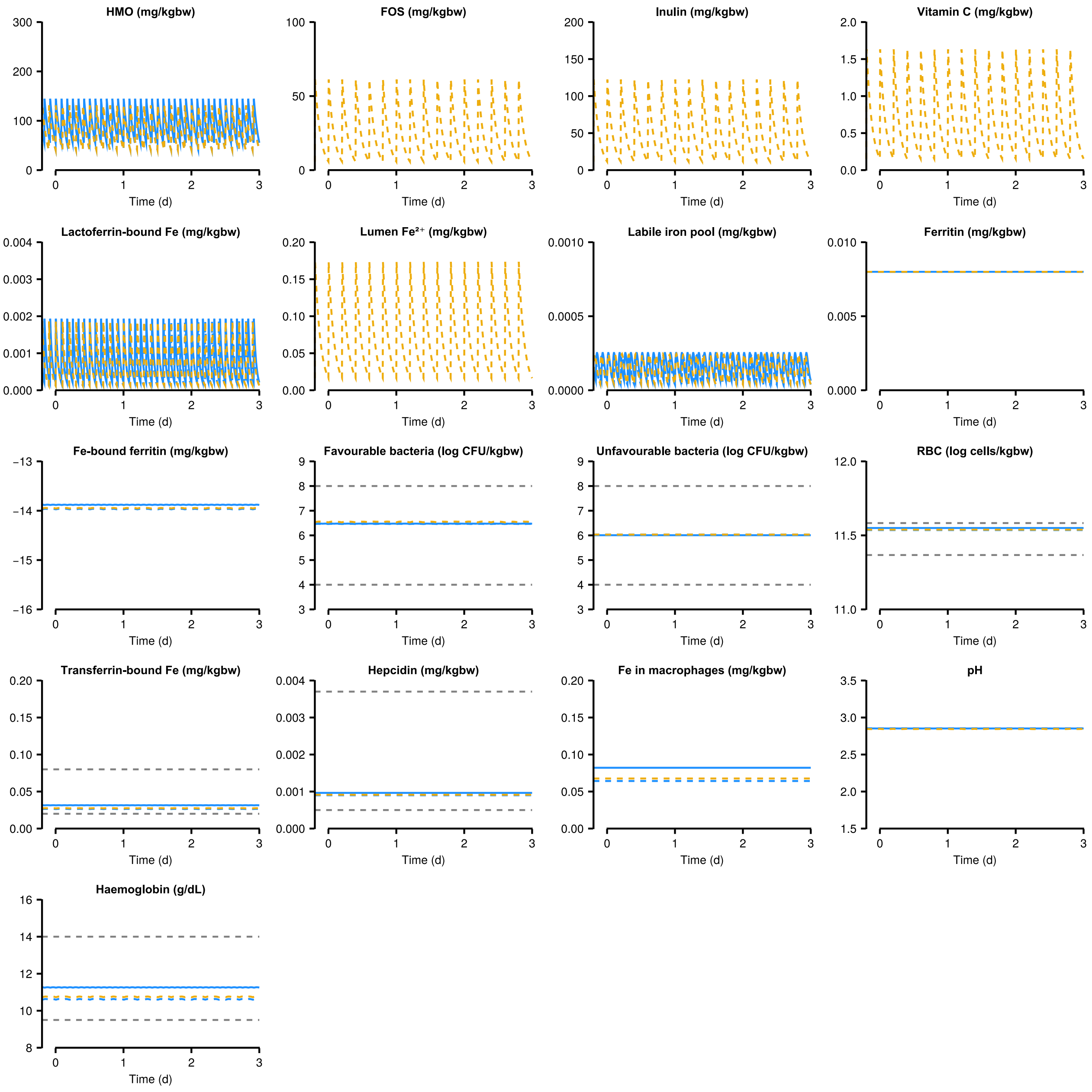


**Fig. S8. Model predictions of all variables in a 4-month-old infant receiving breast milk at deprived frequency and the nutritional solution.**

Steady state dynamics of all model variables in a 4-month-old infant in Fig. S2 receiving breast milk at the frequency of 10 /d (dash, blue) and the effect of nutritional solution (Table S4) in addressing the deprivation (dash, orange). The solid blue line indicates the predictions for the same infant receiving breast milk at the recommended 12 /d. Normal ranges are indicated using grey dashed lines, same as in Fig. S1.

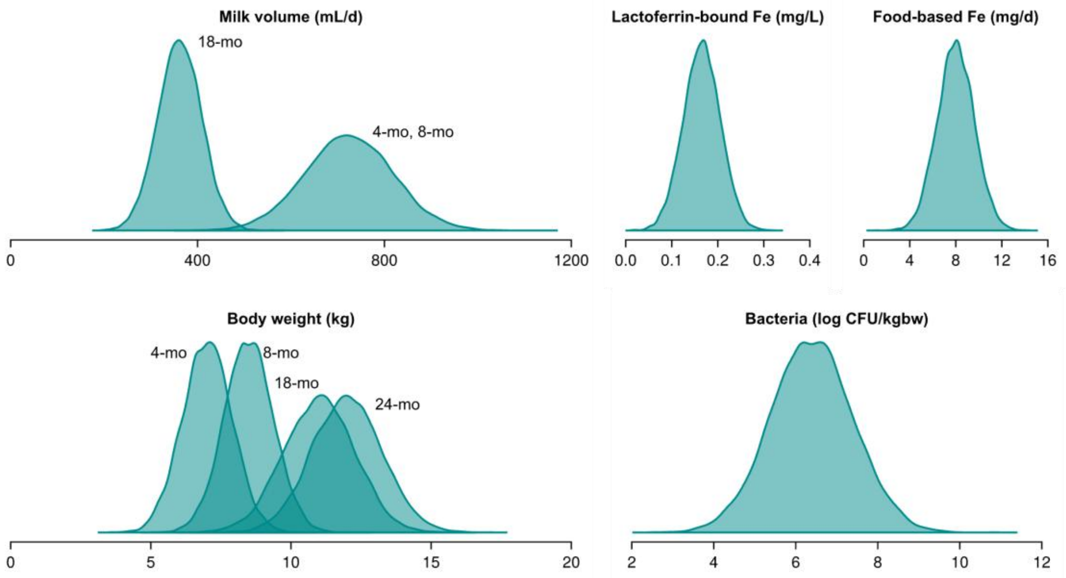


**Fig. S9. Distributions of the quantities indicated in populations of different age groups.**

Sets of samples drawn from these distributions (Table S2) comprise the individuals in calculations involving virtual populations.

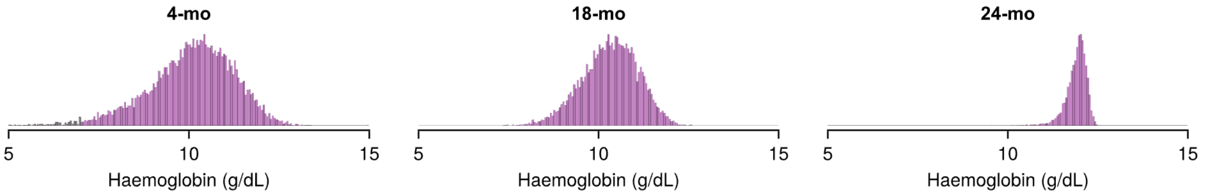


**Fig. S10. Haemoglobin distributions in virtual populations.**

Distribution of haemglobin levels in virtual populations in the three age groups indicated, obtained by sampling parameter combinations (Table S2) to recapitulate the prevalence of mild, moderate, and severe anaemia in the NFHS-5 survey (see Main text).

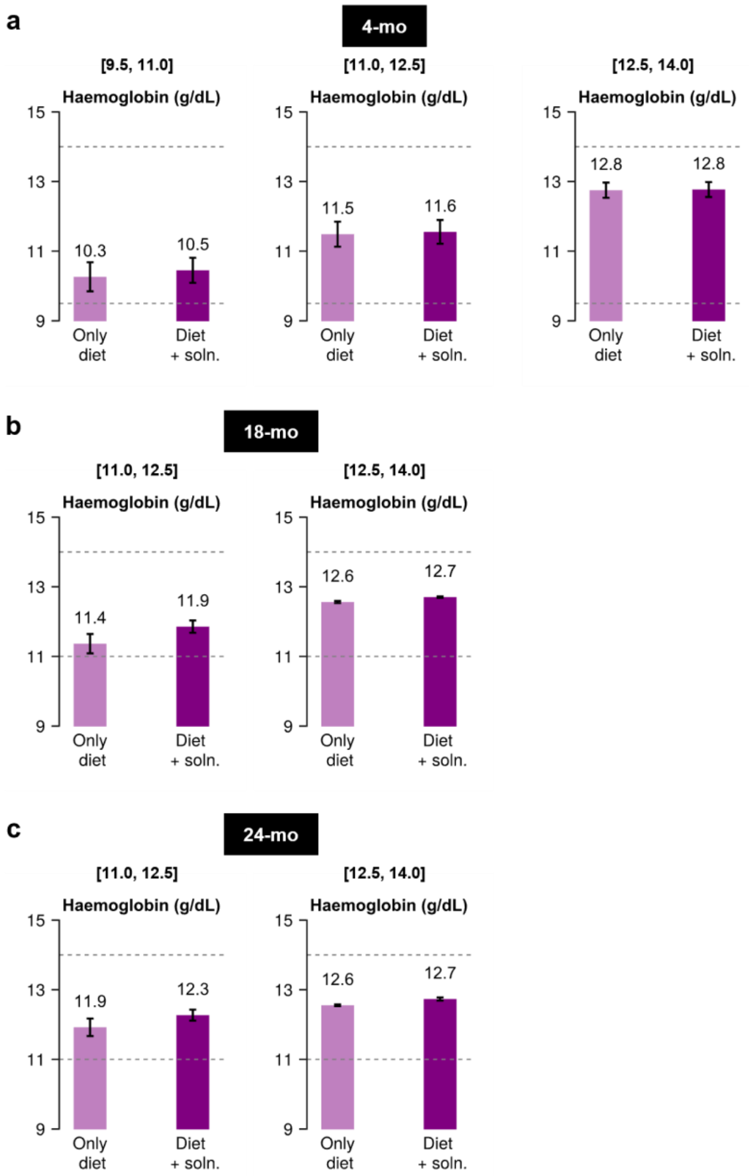


**Fig. S11. Administering nutritional solutions to healthy children.**

Mean haemoglobin levels before (light bar) and after (dark bar) administering age-appropriate nutritional solutions in healthy children from the virtual populations of age 4-mo **(a)**, 18-mo **(b)**, and 24-mo **(c)** (Table S4). The ranges of normal haemoglobin levels in the respective age groups have been subdivided into smaller intervals, indicated above the panels, to better understand the implications of supplementation. Numbers above the bars indicate their heights. Error bars are standard deviations. The horizontal dashed lines indicate the normal ranges (Table S3). Supplementation produces a larger increase in haemoglobin levels in children with lower baseline values while maintaining them in the normal ranges in all subgroups.

### Table S1: Model parameters, values, and their sources

| Parameter | Units | Value | Description |
|---|---|---|---|
| $D_L$ | mg/k-bw | - | Input dose of lactoferrin-bound Fe from breast milk. Bioavailable lactoferrin-bound Fe in mother's milk is 0.165 $\pm$ 0.08 mg/kgbw[5], but the input dosage varies with the amount of breast milk consumed (see Main text) |
| $\tau_{M,i}$ | d | - | Time of $i^{th}$ breast feeding |
| $k_L$ | /d | 15.6 | Absorption rate of lactoferrin-bound Fe[6] |
| $\phi_L$ | /d | 12 | Clearance rate of lactoferrin-bound Fe from intestinal lumen[7] |
| $D_H$ | mg/kgbw | - | Input dose of HMO from breast milk. Breast milk contains 10 g/L of HMO[8], but the input dosage varies with the amount of breast milk consumed (see Main text) |
| $\kappa_H$ | mg/d-CFU | 1.2E-7 | Maximal consumption rate of HMO by bacteria[9] |
| $K_H$ | mg/kgbw | 333.33 | Sensitivity of HMO consumption by favourable bacteria on HMO concentration (assumed) |
| $\phi_H$ | /d | 12 | Clearance rate of HMO from intestinal lumen[7] |
| $\mu_B$ | /d | 7.2 | Maximum growth rate of favourable bacteria[9] |
| $\eta_{B0}$ | CFU/kgbw | 2.5E6 | Basal carrying capacity of favourable bacteria; varies with the child[1] |
| $a_{HB}$ | d | $1/\kappa_H$ | Fold-change in carrying capacity of favourable bacteria with consumption of HMO[10] |

| Parameter | Units | Value | Description |
|---|---|---|---|
| $a_F$ | d | $1.5/\kappa_F$ | Fold-change in carrying capacity of favourable bacteria with consumption of FOS[11] |
| $a_I$ | d | $1/\kappa_I$ | Fold-change in carrying capacity of favourable bacteria with consumption of inulin[11] |
| $\alpha_{BP}\left(=\alpha_{PB}\right)$ | kgbw/mg-d | 1.2E-7 | Competition between favourable and unfavourable bacteria[12] |
| $\mu_P$ | /d | 0.72 | Maximum growth rate of unfavourable bacteria[13] |
| $\eta_{P0}$ | CFU/kgbw | 2.5E6 | Basal carrying capacity of unfavourable bacteria; varies with the child (assumed to be similar to the levels of favorable bacteria) |
| $a_{HP}$ | d | $6/(7\kappa_H)$ | Fold-change in carrying capacity of unfavourable bacteria with consumption of HMO[14] |
| $a_{Fe}$ | cell-d/mg | $2/\kappa_{Fe}$ | Fold-change in carrying capacity of unfavourable bacteria with $Fe^{2+}$ consumption[15] |
| $D_{FeF}$ | mg/kgbw | - | Input dose of Fe from food; varies with food consumed |
| $\tau_{F,j}$ | d | - | Time of $j^{th}$ food intake |
| $D_{FeS}$ | mg/kgbw | - | Input dose of iron from nutritional solution; varies with the solution consumed |
| $\tau_{S,k}$ | d | - | Time of $k^{th}$ nutritional solution dose |
| $c_{max}$ | /d | 0.5 (4-, 18-mo)<br>50 (24-mo) | Maximal $Fe^{3+}\rightarrow Fe^{2+}$ conversion rate[16] |
| $K_{pH}$ | - | 5 | Sensitivity of $Fe^{3+}\rightarrow Fe^{2+}$ conversion to pH (assumed) |

| Parameter | Units | Value | Description |
|---|---|---|---|
| $K_{Vit}$ | mg/kgbw | 0.715 | Sensitivity of $Fe^{3+}$→$Fe^{2+}$ conversion to vitamin C (assumed) |
| $\phi_{Fe3}$ | /d | 12 | Clearance rate of $Fe^{3+}$ from intestinal lumen[7] |
| $\kappa_{Fe}$ | mg/d-CFU | 6E-11 | Maximal consumption rate of $Fe^{2+}$ by unfavourable bacteria (assumed) |
| $K_{Fe}$ | mg/kgbw | 0.406 | Sensitivity of $Fe^{2+}$ consumption by unfavourable bacteria on $Fe^{2+}$ level (assumed) |
| $k_{enter}$ | /d | 30 | Maximal rate of transfer of $Fe^{2+}$ into enterocytes (assumed) |
| $\beta$ | mg/kgbw | 1.6E-16 (4-, 18-mo)<br>3.2E-16 (24-mo) | Sensitivity of $Fe^{2+}$ entry into enterocytes on Fe-bound ferritin level (assumed) |
| $\phi_{Fe2}$ | /d | 12 | Clearance rate of $Fe^{2+}$ from intestinal lumen[7] |
| $\omega$ | - | 3.5 | Constants governing the dependence of intestinal pH on the level of favourable bacteria (assumed) |
| $\nu$ | - | 0.1 | |
| $k_{on}$ | /d | 1 | Unbinding rate of Fe-bound ferritin (assumed) |
| $k_{off}$ | kgbw/d-mg | 1E-8 | Binding rate of Fe and free ferritin (assumed) |
| $k_{utilize}$ | /d | 1E-2 | Utilization of Fe by enterocytes from labile iron pool (assumed) |
| $k_{fpn}$ | /d | 100 | Maximal transport rate of Fe from enterocytes into blood (assumed) |
| $\theta$ | mg/kgbw | 0.0035 | Sensitivity of Fe transport from enterocytes to hepcidin level (assumed) |
| $f_p$ | kgbw/cell | 5E-11 | Fe taken up by a single RBC during its production (assumed) |

| Parameter | Units | Value | Description |
|---|---|---|---|
| $\delta_R$ | /d | 0.00833 | Death rate of RBCs, given their lifespan is 120 d[17] |
| $\gamma$ | mg/kgbw | 1.4E-3 | Sensitivity of Fe release into blood from macrophages to hepcidin level (assumed) |
| $\delta_{TFe}$ | /d | 0.35 (4-mo)<br>0.15 (18-mo)<br>0.12 (24-mo) | Clearance rate of serum transferrin-bound Fe (assumed) |
| $\delta_{MFe}$ | /d | 0.0231 | Death rate of macrophages (assumed) |
| $h$ | mg/d-kgbw | 0.0016 | Maximal hepcidin production rate[18] |
| $K_T$ | mg/kgbw | 0.02 | Sensitivity of hepcidin production to serum transferrin-bound Fe (assumed) |
| $K_p$ | cell/d-kgbw | 2E11 | Sensitivity of hepcidin production to RBC production rate (assumed) |
| $\delta_H$ | /d | 1.0 | Clearance rate of hepcidin[19] |
| $p_{max}$ | cell/d-kgbw | 2.5E10 | Maximal RBC production rate[20] |
| $\epsilon$ | cell/kgbw | 7.5E10 | Sensitivity of RBC production to RBC levels, mediated by erythropoietin (assumed) |
| $\zeta$ | mg/kgbw | 0.015 | Sensitivity of RBC production on serum transferrin-bound Fe level (assumed) |
| $k_1$ | - | 15.4 (4-mo)<br>16.625 (8-, 18-, and 24-mo) | Constants of the function relating the steady state transferrin-bound Fe in serum and haemoglobin levels (estimated) |
| $k_2$ | mg/kgbw | 0.008 (4-mo)<br>0.015 (8-, 18-, and 24-mo) | |
| $m$ | - | 1 | Hill coefficients (assumed) |
| $n$ | - | 2 | |
| $q$ | - | 1 | |

| Parameter | Units | Value | Description |
|---|---|---|---|
| $D_F$ | mg/kgbw | - | Input dose of FOS from nutritional solution; varies with the child and the corresponding solution used. |
| $\kappa_F$ | mg/d-CFU | 1.2E-7 | Rate of consumption of FOS by favourable bacteria (assumed) |
| $K_F$ | mg/kgbw | 333.33 | Sensitivity of FOS consumption by favourable bacteria to FOS concentration (assumed) |
| $\phi_F$ | /d | 12 | Clearance rate of FOS from intestinal lumen[7] |
| $D_I$ | mg/kgbw | - | Input dose of inulin from nutritional solution; varies with the child and the corresponding solution used. |
| $\kappa_I$ | mg/d-CFU | 6E-8 | Rate of consumption of inulin by favourable bacteria (assumed) |
| $K_I$ | mg/kgbw | 333.33 | Sensitivity of inulin consumption by favourable bacteria to inulin concentration (assumed) |
| $\phi_I$ | /d | 12 | Clearance rate of inulin from intestinal lumen[7] |
| $D_V$ | mg/kgbw | - | Input dose of vitamin C from nutritional solution; varies with the and the corresponding solution used. |
| $\phi_V$ | /d | 12 | Clearance rate of vitamin C from intestinal lumen[7] |

Note:

- Since 8-month-old children would be physiologically closer to the 4-month-olds than the others, we assumed all the parameter values except for $k_1$ and $k_2$ of the former identical to that of the latter.
- 'kgbw' is kilogram body weight

### Table S2: Physiological characteristics

| Description | Ranges | Source |
|---|---|---|
| ***Children*** | | |
| Body weight (kg) | 4-mo: 5.5 – 9<br>8-mo: 7 – 10.5<br>18-mo: 9 – 14<br>24-mo: 10 – 15 | WHO[21] |
| Fe RDA (mg/d) | 4-mo: NA<br>8-mo: 6<br>18-, 24-mo: 8 | ICMR-NIN recommendations[22] |
| Maximal favorable bacteria levels ($log_{10}$ CFU/kgbw) | 6.40 $\pm$ 2 | Mean value from Table S1; variation assumed |
| Maximal unfavorable bacteria levels ($log_{10}$ CFU/kgbw) | 6.40 $\pm$ 2 | Mean value from Table S1; variation assumed |
| ***Mothers*** | | |
| Breast milk volume fed (mL/d) | 4-, 8-mo: 724 $\pm$ 184<br>18-mo: 372 $\pm$ 184<br>24-mo: NA | [23] |
| Bioavailable lactoferrin-bound Fe (mg/L) | 0.165 $\pm$ 0.08 | [5] |

Note:

- The recommended frequency of breastfeeding is 12 /d, and the feed is assumed to be equally split between the 12 doses.
- We assume that the dependence on breast milk weans off by 24-month age and the child from then on completely depends on food diet for iron.

### Table S3: Clinical ranges

| Description | Ranges | Source |
|---|---|---|
| Haemoglobin (g/dL) | (4-mo)<br>Normal: 9.5 – 14<br>Mild anaemia: 9.0 – 9.5<br><br>(8-, 18-, 24-mo)<br>Normal: 11 – 14<br>Mild anaemia: 10 – 10.9<br>Moderate anaemia: 7 – 9.9<br>Severe anaemia: <7 | [4] |
| Serum transferrin–bound Fe (mg/kgbw) | Normal: 0.02 – 0.08 | [2] |
| Gut bacteria (cell/kgbw) | Normal: 1E4 – 1E8 | [1] |

### Table S4: Details of the nutritional solutions

| Description | Age of the child (age group pertaining to the supplement) | Composition per dose | Recommended number of doses per day |
|---|---|---|---|
| DS-2 | 2-mo (0–6 months) | FOS: 0.405 g<br>Inulin: 0.81 g<br>Fe: 1.15 mg<br>Vitamin C: 10.8 mg | 5 |
| Dexolac 3 | 18-mo (12–18 months) | Fe: 1.62 mg<br>Vitamin C: 10.8 mg | 3 |
| Dexolac 4 | 24-mo (18–24 months) | Fe: 1.62 mg<br>Vitamin C: 10.8 mg | 2 |

**Table S5: Change in haemoglobin levels with intervention as reported in a clinical trial and comparing them with model predictions**

| **Reference** | Pasricha et al., *N Engl J Med* 2021[24] | |
|---|---|---|
| Study period | July 2017 – February 2019 (3 months intervention, follow-up: 9 months post-intervention) in rural Bangladesh | |
| Sample size | Group-1: 1101 on iron syrup<br>Group-2: 1099 on micronutrient powder (MNP)<br>Group-3: 1100 on placebo | |
| Intervention details | Group-1: Iron syrup, containing Fe as $Fe^{2+}$ salt, given daily to provide 12.5 mg elemental Fe in a single dose<br>Group-2: MNP, containing Fe as $Fe^{2+}$ salt, given daily to provide 12.5 mg elemental Fe in a single dose<br>Group-3: Placebo | |
| **Status of the groups** | **Study observation** | **Model prediction** |
| Baseline haemoglobin levels (g/dL) | Group-1: 11.0 ± 1.0 | 10.99 ± 0.87 |
| | Group-2: 11.0 ± 1.0 | 11.0 ± 1.0 |
| | Group-3: 11.0 ± 1.0 | 11.02 ± 0.88 |
| Haemoglobin levels after intervention (g/dL) | Group-1: 11.55 ± 1.37 | 11.41 ± 0.84 |
| | Group-2: 11.47 ± 1.37 | 11.42 ± 0.87 |
| | Group-3: 10.87 ± 1.29 | 11.0 ± 0.88 |
| Prevalence of anaemia (%) | Group-1:<br>Before – 46.2<br>After – 23.4 | Group-1:<br>Before – 47.8<br>After – 26.4 |
| | Group-2:<br>Before – 43.8<br>After – 25.6 | Group-2:<br>Before – 49.7<br>After – 28.5 |
| | Group-3:<br>Before – 44.4<br>After – 49.2 | Group-3:<br>Before – 49.6<br>After – 49.6 |
